# Superspreading of SARS-CoV-2: a systematic review and meta-analysis of event attack rates and individual transmission patterns

**DOI:** 10.1101/2024.01.25.24301669

**Authors:** Clifton D. McKee, Emma X. Yu, Andrés Garcia, Jules Jackson, Aybüke Koyuncu, Sophie Rose, Andrew S. Azman, Katie Lobner, Emma Sacks, Maria D. Van Kerkhove, Emily S. Gurley

## Abstract

SARS-CoV-2 superspreading occurs when transmission is highly efficient and/or an individual infects many others, contributing to rapid spread. To better quantify heterogeneity in SARS-CoV-2 transmission, particularly superspreading, we performed a systematic review of transmission events with data on secondary attack rates or contact tracing of individual index cases published before September 2021, prior to emergence of variants of concern and widespread vaccination. We reviewed 592 distinct events and 9,883 index cases from 491 papers. Meta-analysis of secondary attack rates identified substantial heterogeneity across 12 chosen event types/settings, with the highest transmission (25–35%) in co-living situations including households, nursing homes, and other congregate housing. Among index cases, 67% produced zero secondary cases and only 3% (287) infected >5 secondary cases (“superspreaders”). Index case demographic data was limited, with only 55% of individuals reporting age, sex, symptoms, real-time PCR cycle threshold values, or total contacts. With the data available, we identified a higher percentage of superspreaders among symptomatic individuals, individuals aged 49–64 years, and individuals with over 100 total contacts. Addressing gaps in reporting on transmission events and contact tracing in the literature is needed to properly explain heterogeneity in transmission and facilitate control efforts for SARS-CoV-2 and other infections.

## INTRODUCTION

Following the emergence of SARS-CoV-2 in late 2019, the virus spread worldwide, resulting in the coronavirus disease (COVID-19) pandemic [1]. Understanding drivers of SARS-CoV-2 transmission was crucial for formulating control measures, especially prior to the development of vaccines. Early in the pandemic, heterogeneity in transmission, particularly superspreading, was investigated because of its ability to cause large outbreaks [2–4].

Superspreading involves two distinct but non-mutually exclusive phenomena: a setting where many people become infected due to an environment conducive to transmission (e.g., crowded indoor settings), and individuals who are outliers in the number of secondary cases they infect, due to biological heterogeneity in infectiousness and/or engagement in high-risk behaviors [5,6]. Superspreading has been observed in several other viral infections, including SARS-CoV, MERS-CoV, Nipah, Ebola, and measles [7–12]. With SARS-CoV-2, both forms of superspreading garnered considerable attention in the literature. For example, over 140 individuals were infected during a Christmas event in Belgium in December 2020, causing over 26 deaths [13]. Likewise, one individual infected dozens of people during a choir practice in Washington, USA, in March 2020 [14].

Because superspreading events contributed substantially to local and global SARS-CoV-2 transmission [15], public health interventions were enacted to reduce their risk of occurrence. These interventions included school closures, limitations on indoor gatherings, and restrictions on visiting hospitalized patients or long-term care facilities. Many of these policies were based on limited data from early in the pandemic. Moreover, published systematic reviews and modeling of SARS-CoV-2 superspreading from this period were limited in scope and did little to disaggregate this phenomenon into the distinct contributions of environment and individual characteristics. For example, studies of setting-specific transmission rates have focused on household and healthcare transmission or geographic and temporal trends [2,16–19], but did not address transmission heterogeneity across other social settings. Previous meta-analyses of individual-level superspreading included only a small number of papers (<26) that calculated overdispersion in transmission, missing the majority of published transmission trees and capturing data primarily from Asia [7,8]. Early investigations of individual-level characteristics related to superspreading were also limited by incomplete contact tracing [20,21] and a focus on clinical over demographic characteristics [20]. A more complete summary of superspreading is needed to understand the scale of transmission heterogeneity across settings and identify causes of individual heterogeneity.

The objective of this review was to summarize global heterogeneity in SARS-CoV-2 transmission events prior to widespread vaccination and the role of environmental and individual factors in superspreading. Specifically, this review aimed to identify: 1) the amount of variation in attack rates across studies and events, 2) which settings had the highest attack rates, 3) the individual offspring distribution for SARS-CoV-2, and 4) the characteristics of superspreading individuals.

## METHODS

### Literature search and data extraction

We conducted this systematic review and meta-analysis according to the Preferred Reporting Items for Systematic Reviews and Meta-Analyses (PRISMA) 2020 statement [22]; see Appendix 1 for the PRISMA checklist. We included all studies of SARS-CoV-2 that contained data on: 1) transmission chains; 2) numbers of index cases, contacts, and infected contacts; 3) numbers of index cases and infected contacts; or 4) secondary attack rates, i.e., number of infected contacts divided by number of contacts. We excluded studies that were not about humans. A clinical informationist searched PubMed, the WHO COVID database, the I Love Evidence COVID database, and Embase on 9 September 2021. No restrictions on language or start date were applied. Results were imported into EndNote X9 (Clarivate, London, UK) where duplicates with exact matches in the author, year, and title fields were removed. Team members screened titles and abstracts and performed full text review in Covidence (Veritas Health Innovation, Melbourne, Australia).

We extracted data using a pre-designed, study-specific spreadsheet, collecting information on paper metadata and target variables for two outcomes: transmission events and individual index cases (Table 1). Events were defined as discrete transmission events where secondary attack rates for defined groups of people could be calculated as the number of infected cases divided by the total number of exposed individuals. This definition of secondary attack rates includes both clinical and subclinical infections in some studies. Due to the limited details published in the literature, we did not attempt to distinguish events associated with individual transmission chains from a single source (potentially with confirmatory sequencing data) from events that aggregated multiple transmission chains together. In lieu of this distinction, we separated events into different settings and by the duration of the event (i.e., exposure window, in days) reported in each paper. Twelve event types were chosen to classify each event/setting described in a paper (Table 2). To describe individual contributions to transmission, we extracted data on index cases for whom contacts were followed to identify secondary transmission. We only entered data from papers where it was clear from the methods that contact tracing was done for at least one week to capture secondary transmission from individual index cases. For studies that did not report SARS-CoV-2 variants, we imputed the dominant variant from CoVariants data for the country and time period of interest [23]. See the Supplementary Material for additional details on the identification of papers, data extraction (Supplementary Tables S2–S3), and assessment of study bias.

**Table 1.** Description of variables extracted from papers in the systematic review of SARS-CoV-2 superspreading from December 2019 to July 2021.

|  |  |
| --- | --- |
| All papers | Title<br>Author(s)<br>Publication year, volume, and issue<br>Journal<br>Study location(s)<br>Country<br>Administrative unit(s): state/province, county, city<br>General study time period (e.g., start and end year/month of data collection)<br>Diagnostic testing method (PCR, serology, rapid antigen tests, symptom diagnosis, or mixed)<br>Variant name<br>Any reported prevention measures implemented in the event (e.g., masking, social distancing)<br>Number of exposed people with reported demographic characteristics (age, sex) and vaccination status |
| Events | Type of event/setting (e.g. nursing home residents, household transmission study, or school)<br>Start and end date of event |
| Index case | Demographic characteristics (age, sex, occupation)<br>Age group: $\leq 4$ years, 5–12 years, 13–18 years, 19–24 years, 25–48 years, 49–64 years, $\geq 65$ years<br>Symptom onset (if applicable) and diagnosis dates<br>Symptoms (text descriptions or presence/absence)<br>Real-time PCR cycle threshold (Ct) value |
|  | • Specimen type<br>• Clinical outcome (if applicable)<br>• Setting of contact (e.g., work, social, and school) |

**Table 2.** Types of SARS-CoV-2 secondary transmission events occurring between December 2019 and August 2021 reported in the literature. Heterogeneity across event types was assessed based on the variance and interquartile range of secondary attack rates. Outlier events were identified for each event type as events that exceeded the estimated upper confidence interval of the meta-analysis estimated SAR for that event type or were greater than 50%.

| Event type | Description of outbreak location | N | Minimum and maximum secondary attack rate |
| --- | --- | --- | --- |
| 1 | cruise ship or other densely populated watercraft (e.g., fishing vessel, aircraft carrier) | 16 | 0.02, 1 |
| 2 | transport mode other than ships (e.g., airplane, train, car) | 20 | 0, 0.4 |
| 3 | households, defined as co-living individuals or close contacts who always meet each other but possibly not living together (e.g., couples in romantic relationship) | 115 | 0, 1 |
| 4 | hospital or healthcare facility, including patients, healthcare workers, and nursing home workers (if worker data was provided separately from nursing home residents) | 89 | 0, 0.46 |
| 5 | workplace (e.g., office), including correctional officers and teachers and staff at schools | 51 | 0, 0.86 |
| 6 | school (data on students only) | 32 | 0, 0.54 |
| 7 | public social venue (e.g., bar, concert, sporting event) | 39 | 0, 1 |
| 8 | private social event with members of multiple households (e.g., dinner with neighbors or extended family) | 12 | 0.01, 0.81 |
| 9 | shopping (activities in shops, markets, and department stores) | 2 | 0, 0 |
| 10 | nursing home or long-term care facility (residents only or residents and healthcare workers if not described separately in the paper) | 41 | 0, 0.84 |
| 11 | congregate housing other than nursing home or long-term care facility<br>(e.g., homeless shelter, prison, summer camp) | 84 | 0, 1 |
| 12 | mixed locations, included any combination of the above but not<br>described separately in the paper | 91 | 0, 1 |

### Statistical analyses

To characterize the type and quality of information that we were able to extract about transmission events, we performed a descriptive analysis of event data including the number of each chosen event type, starting year of the data, focal countries, diagnostic methods, event duration, and level of missingness for all variables. Because not all individuals potentially exposed during an event were tested in each study, secondary attack rates for individual events were calculated separately using the total number of exposed individuals or the total number tested. If either of these quantities were missing, the value was imputed based on the value present (i.e., assuming the number tested was equal to the number exposed or vice versa). Sensitivity of results to this choice of denominator was assessed in the meta-analysis of events (see Supplementary Material).

To describe the amount of variation in attack rates across studies and events and to identify which settings had the highest SARS-CoV-2 attack rates, a meta-analysis was performed on secondary attack rates across event types using the *metafor* package in R v4.2.2 [24]. We converted secondary attack rates for each event to Freeman-Tukey double arcsine transformed proportions [25] and calculated the sampling variance. We fit a hierarchical model with a nested random effect for event within study and no fixed effects to assess the heterogeneity in secondary attack rates attributable to these factors using restricted maximum likelihood. We calculated *I^2^*, the percentage of variance attributable to true heterogeneity, for each random effect [26] and used Cochran’s *Q* test to test if estimated heterogeneity in secondary attack rates was greater than expected from the sampling error alone. We then fit additional mixed-effects models that included the same random effects but also event type and event duration as fixed effects. Cochran’s *Q* was performed on these model to assess whether residual heterogeneity in secondary attack rates was greater than expected after accounting for sampling error and fixed effects. Fitted coefficients and 95% confidence intervals (CI) from meta-analysis were back-transformed to proportions using the geometric mean of the tested individuals across all studies in each event type [25]. These back-transformed proportions are referred to as “meta-analysis estimated secondary attack rates” or “meta-analysis estimated mean attack rates” in the text and figures. For comparison with meta-analysis estimates, we also calculated the median secondary attack rate and interquartile range across events for each chosen event type.

To characterize the individual offspring distribution for SARS-CoV-2, the overall distribution of secondary cases generated by each identified index cases was fit to a negative binomial distribution, following Lloyd-Smith et al. [11]. We estimated the percentile of index cases producing 80% of all secondary infections using a formula and code from Endo et al. [27].

Our last aim for the study was to identify recognizable characteristics of superspreading individuals. Based on the availability of demographic characteristics and other features of index cases in the literature, we examined differences in distributions of secondary cases produced by index cases according to sex, presence/absence of symptoms, age, real-time PCR cycle threshold (Ct) value, and the total number of contacts each index case had. Additional statistical tests compared these listed factors between “superspreaders” (index cases with >5 secondary cases, following Adam et al. [3]) and “non-superspreaders” (index cases with ≤5 secondary cases): Chi-square tests of proportions to compare the proportion of women, the proportion of symptomatic cases, and proportion of adults or across age bins; Student’s t-tests to compare mean age and Ct value; and a Kruskal-Wallis test to compare the highly skewed distributions of total contacts among index cases. All statistical tests used α = 0.05 as the statistical significance threshold to identify whether superspreaders were overrepresented among certain demographic groups.

## RESULTS

### Study selection

We identified 13,632 articles from the four databases searched, representing 8,339 unique references (Figure 1). Of these, we excluded 7,358 records during the abstract review. For the 981 records that underwent full text review, we excluded 384 records that were reviews or letters to the editor without data, contained no data on our variables of interest, or were duplicate records (preprints, true duplicates, or duplicated datasets). A total of 598 papers were assessed for eligibility for data extraction and a further 107 papers were excluded that did not contain sufficient data on our outcome variables of interest or were duplicates (Figure 1). We extracted data from 491 studies: 232 studies provided event data only, 195 studies provided individual index case data only, and 64 studies provided both data types, yielding evidence from 592 distinct events and 9,883 index cases. The 491 analyzed studies were from 67 countries, with most from China (26%), the USA (17%), and South Korea (5%) (Supplementary Figure S1A).

**Figure 1.**
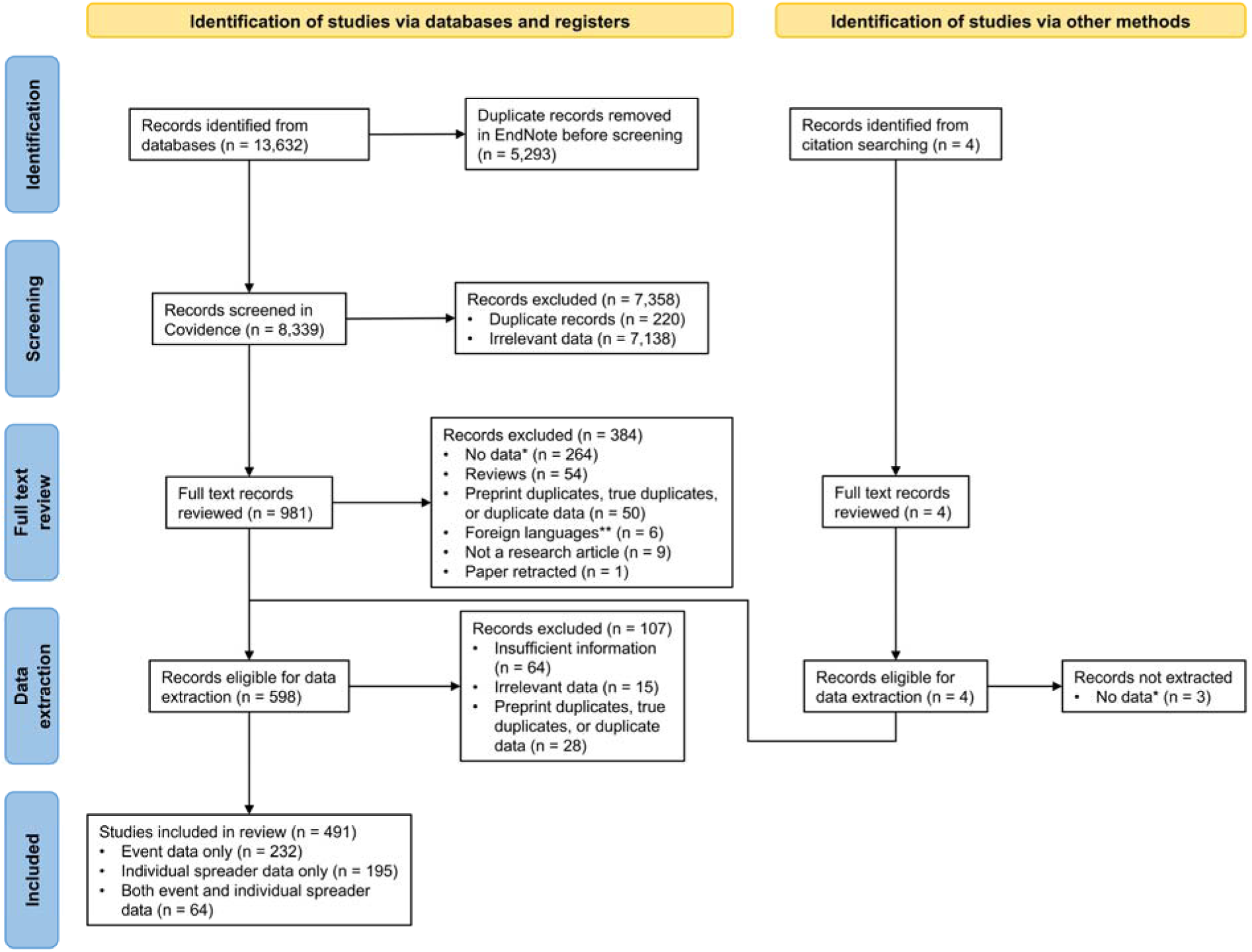
PRISMA flow diagram for the systematic review and meta-analysis of SARS-CoV-2 superspreading reported in the published literature. *There were 4 types of data that we sought to include: 1) transmission chain; 2) number of index cases, number of contacts, and number of infected contacts; 3) number of index cases and number of infected contacts; or 4) secondary attack rate. **Languages other than Spanish, Chinese, French, Turkish, German, and Portuguese.

Although our search included two-thirds of 2021, nearly all studies covered data from 2020 (94% of events, 99% of index case symptom onset or positive test dates).

### Characteristics of events

Descriptive analyses were used to characterize the type and quality of information about transmission events present in the literature. Event data were most commonly from the USA (27%), China (15%), the UK (8%), and South Korea (6%) (Supplementary Figure S1B).

Published papers were missing information on many variables that we aimed to extract about events (Supplementary Figure S2A). Of the 46 target data fields from articles about events, 17 had high data completeness (>80%), including those for the study and event metadata, event description, time period of the event (describing the start and end dates of exposure), location of the event (country and state/province or city), and number of exposed individuals and secondary cases (Supplementary Table S3). Event durations were highly skewed, with a median duration of 34 days and an interquartile range of 13–60 days (Supplementary Figure S3). Studies used a variety of diagnostic methods to identify SARS-CoV-2 cases, though PCR was the dominant method (Supplementary Figure S4A). Other approaches included antigen tests, retrospective case identification by serology, diagnosis via symptoms or chest tomography in early papers, or a mixture of approaches. Because most studies covered events prior to emergence of variants, most events (N = 532, 90%) likely involved only wild-type/ancestral SARS-CoV-2, while 14 events involved Alpha, six Beta, eight Delta, and 31 likely included a mixture of variants (e.g., during periods of variant emergence and replacement of the dominant variant).

### Heterogeneity in event secondary attack rates

Meta-analysis of secondary attack rates was performed to describe variation in attack rates across studies and events and to identify which settings had the highest attack rates. Secondary attack rates varied substantially within and among event types (Figure 2). Interquartile ranges of attack rates were lower for transport (0–11%), hospital/healthcare (1–20%), and mixed events (3–12%), whereas congregate housing (9–63%), households (15–60%), social venues (8–53%), and cruise ships (9–41%) had higher heterogeneity, with some events reporting attack rates of 100% (Table 2). Meta-analysis of secondary attack rates including a nested random effect for event within study detected significant heterogeneity in secondary attack rates (*I^2^* = 99%, Cochran’s *Q_E,_*_591_ = 141,765, *P* < 0.0001). The random effect for study accounted for most of the heterogeneity (*I^2^* = 58%), followed by event nested within study (*I^2^* = 41%). Addition of a fixed effect for event type to the model indicated that secondary attack rates varied significantly across event types (Cochran’s *Q_M,_*_11_ = 122, *P* < 0.0001). Meta-analysis estimated mean attack rates were lowest for shopping (0%), hospitals and healthcare (6%), transportation other than cruise ships (9%), and schools (11%) (Figure 2). Comparatively, estimated mean attack rates were two to three times higher (25–35%) in nursing homes, cruise ships, households, and other congregate housing settings (e.g., homeless shelters, prisons).

**Figure 2.**
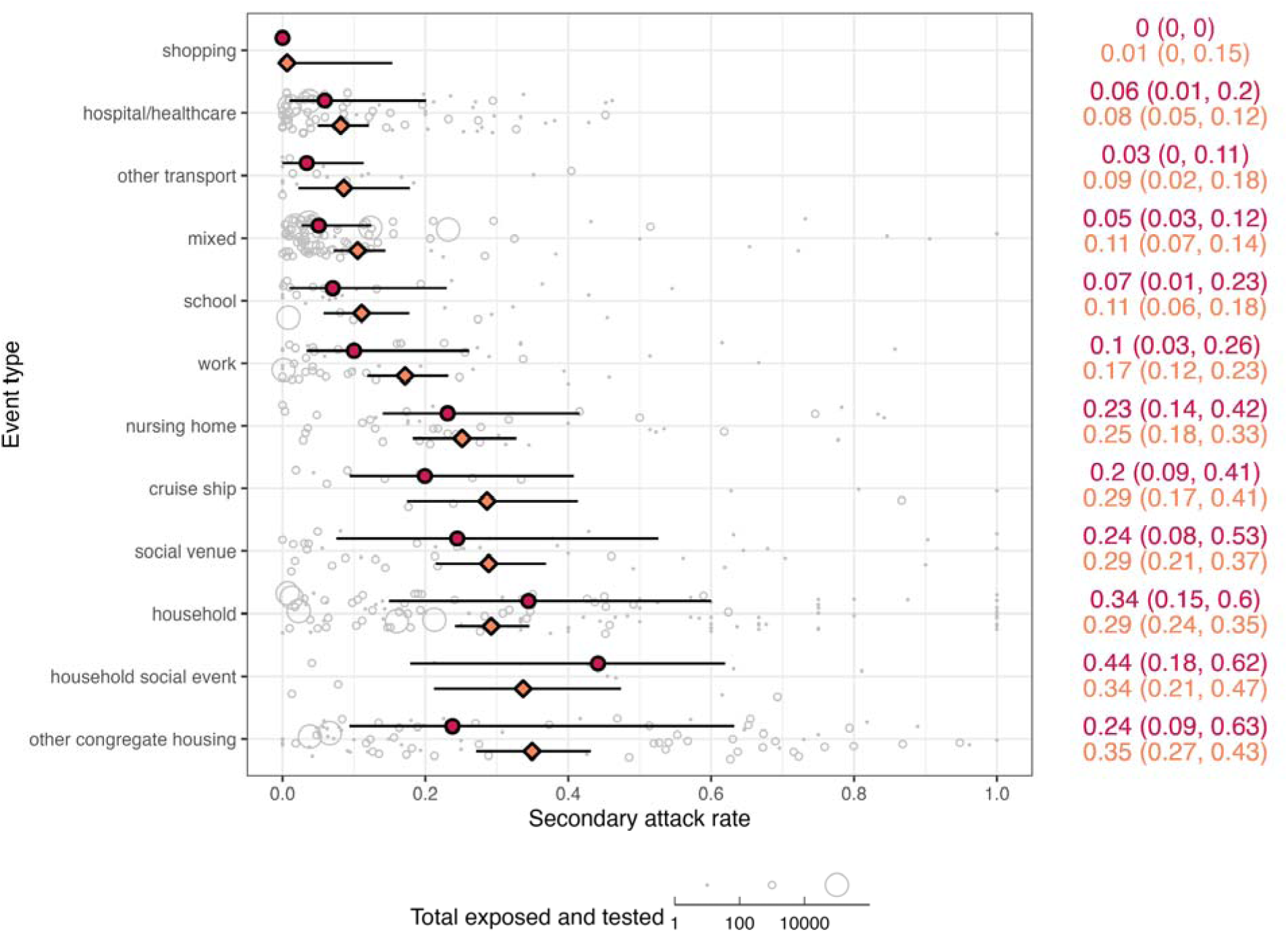
SARS-CoV-2 secondary attack rates across 12 event types occurring between December 2019 and August 2021 reported in the literature across 592 events from 296 studies. Individual event data secondary attack rates are shown as grey bubbles, varying in size according to the total number of individuals exposed and tested from the event. Median secondary attack rate for each event type is shown as red circle with a line representing the interquartile range; values are in red on the right side of the figure. Meta-analysis estimated secondary attack rate for each event type is shown as an orange diamond with a line representing the estimated 95% confidence interval; values are in orange on the right side of the figure. Event types were ranked by increasing estimated mean secondary attack rate along the left axis.

Models including event duration and an interaction term between event type and event duration as additional fixed effects found similar levels of heterogeneity (Cochran’s *Q_M,_*_23_ = 135, *P* < 0.0001) and identified a common trend of decreasing attack rates with longer event durations across different event types, with the exception of cruise ships and shopping (Supplementary Figure S5).

### Characteristics of individual index cases

Descriptive analyses were also used to characterize the type and quality of information about individual index cases found in published studies. Individual index case data with offspring distributions overwhelmingly came from China (36%) and India (35%) (Supplementary Figure S1C). Index case data exhibited higher missingness compared to events (Supplementary Figure S2B): of the 74 data fields that we extracted for individual index cases, the highest completeness (>60%) was seen for study and index case numbers, location of the index case (country and state/province or city), total number of contacts infected, method of testing for the index case and contacts, and SARS-CoV-2 variant (Supplementary Table S4). We identified five key characteristics of index cases that could be related to superspreading, though most of these were also missing from the published literature: 46% of cases included data on age, 48% on sex, 10% on presence/absence of symptoms, 6% on total number of contacts, and only 2% had Ct values reported. A total of 5,437 index cases (55%) contained data on at least one of these five variables. Diagnostic methods for identification of individual index cases and their associated secondary cases were only reported in 61% of cases, with PCR as the primary approach (Supplementary Figure S4B,C). The majority of index cases (N = 8,565, 87%) were assumed to be infected with wild-type SARS-CoV-2 based on location and timing of the study or test confirmation date. A mixture of variants was likely in 1,282 cases (13%), while one index case was reported with Alpha, two Beta, 11 Delta, and 22 Epsilon.

### Heterogeneity in transmission across individual index cases

A third goal of this analysis was to describe the individual offspring distribution for SARS-CoV-2 based on reported index cases. Most index cases (67%) did not transmit SARS-CoV-2 to another person and 17% transmitted to only one other individual (Figure 3). There were 287 “superspreaders” with >5 contacts infected, representing 3% of index cases from the included studies. The distribution of secondary infections fit a negative binomial distribution with a mean of 0.88 (CI: 0.84–0.92) and a dispersion parameter *k* of 0.27 (CI: 0.25–0.28). Using the formula from Endo et al. [27] and the estimated mean and *k* for the negative binomial distribution, the top 17% most infectious index cases would be expected to generate 80% of all secondary cases.

**Figure 3.**
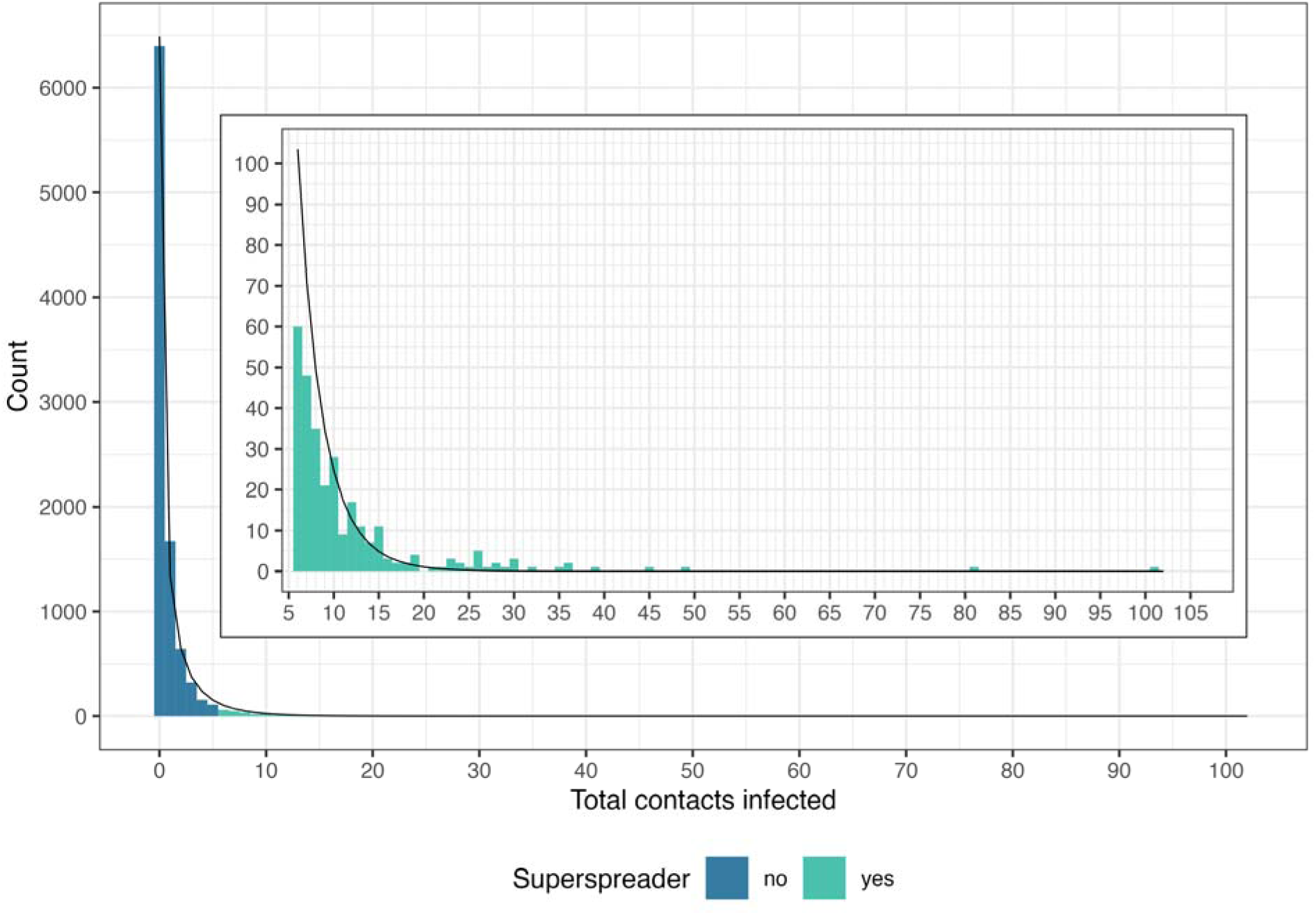
Distribution of secondary contacts infected by individual index cases (N = 9,591) for SARS-CoV-2 cases occurring between December 2019 and July 2021 reported in 259 studies. The black line shows the fit of the distribution to the expected negative binomial distribution. The inset shows a portion of the same data to highlight the distribution of superspreaders (index cases with >5 secondary cases).

### Qualities of superspreaders

Finally, our analysis sought to identify qualities of index cases that were associated with being a superspreader (index cases with >5 secondary cases) compared to non-superspreaders (Table 3). The proportion of index cases with reported symptoms was higher in superspreaders (89%) than non-superspreaders (76%; ^2^ = 5.4, *P* = 0.02). Superspreaders had more than two times the mean number of contacts (79) compared to non-superspreaders (36; χ^2^ = 56.6, *P* < 0.0001). Adults also made up a greater proportion of superspreaders (99%) than non-superspreaders (84%; ^2^ = 14.1, *P* < 0.0001). Index cases over 25 years of age were overrepresented among superspreaders and no superspreaders 12 years of age and under were reported (Figure 4). When age was analyzed as a continuous variable, the number of contacts infected and the frequency of superspreaders increased with age, up to around 60 years of age (Supplementary Figure S6). No significant differences by sex or Ct values were observed (Table 3). However, two adult male index cases produced the highest number of secondary infections, infecting 81 of their 104 contacts and 101 of their 300 contacts, respectively. The former was a lecturer in Tonghua, China [28] and the latter a fitness instructor in Hong Kong, China [29].

**Figure 4.**
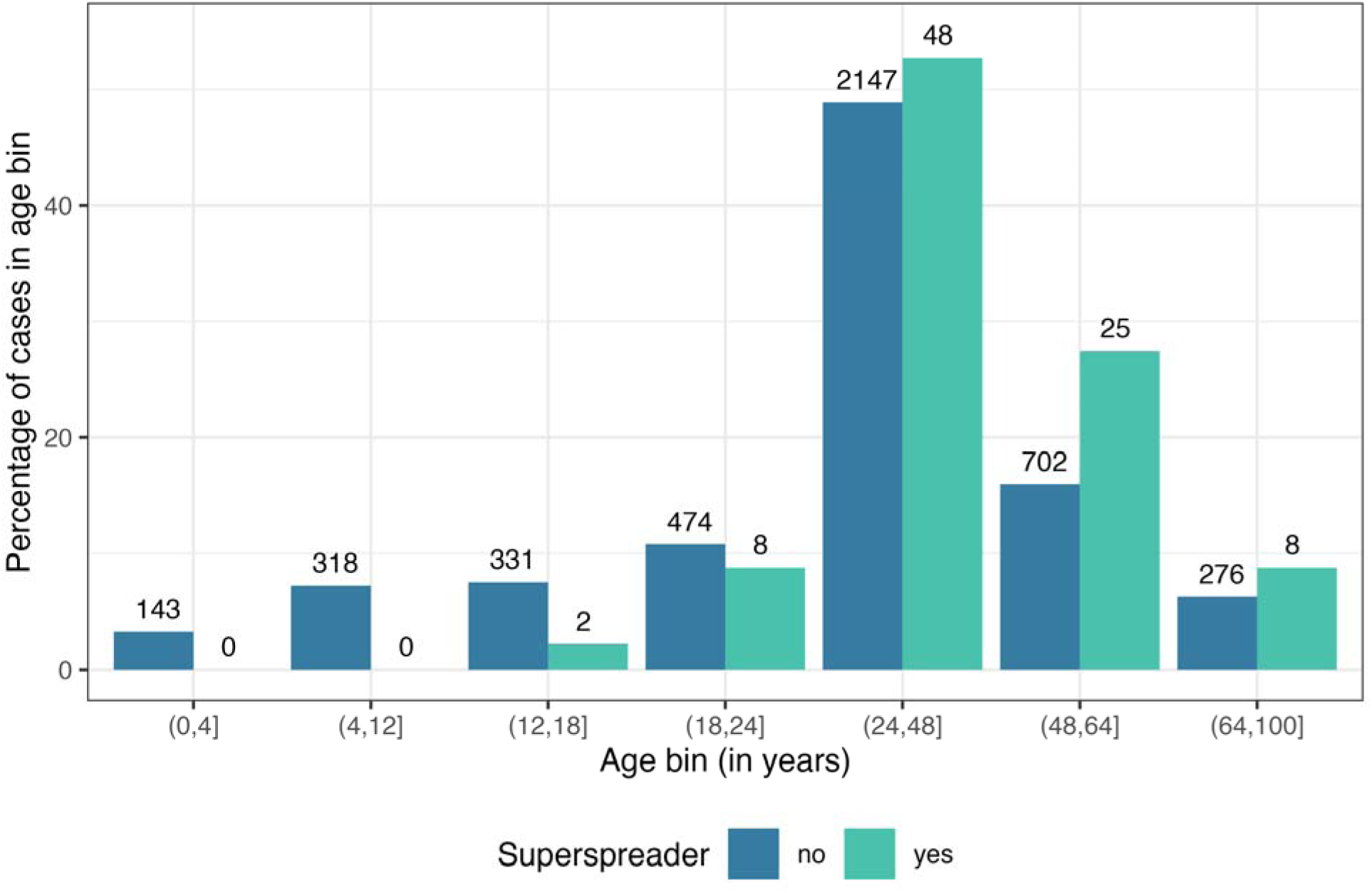
Comparison of the age distribution of superspreading index cases. The bars show the percentage of individuals within an age bin across superspreaders (index cases with >5 secondary cases) and non-superspreaders. Numbers above the bars display the raw totals and percentages are shown in Table 3.

**Table 3.** Statistical comparisons of SARS-CoV-2 superspreaders to non-superspreaders based on features reported in the literature in 259 studies for cases occurring between December 2019 and July 2021.

| Feature of comparison | Percentage or estimated mean for non-superspreaders (total observations) | Percentage or estimated mean for superspreaders (total observations) | Statistical test results |
| --- | --- | --- | --- |
| Female | 40% (N = 4,543) | 38% (N = 102) | $\chi^2_1 = 0.09, P = 0.76$ |
| Presence of symptoms (symptomatic) | 76% (N = 841) | 89% (N = 70) | $\chi^2_1 = 5.4, P = 0.02$ |
| Age (in bins) | (N = 4,391) | (N = 91) | $\chi^2_6 = 21.7, P = 0.001$ |
| ≤4 years | 3% | 0% |  |
| 5–12 years | 7% | 0% |  |
| 13–18 years | 8% | 2% |  |
| 19–24 years | 11% | 9% |  |
| 25–48 years | 49% | 53% |  |
| 49–64 years | 16% | 27% |  |
| ≥65 years | 6% | 9% |  |
| Age (≥18 years) | 84% (N = 4,391) | 99% (N = 91) | $\chi^2_1 = 14.1, P < 0.0001$ |
| Age (in years) | 34.8 (N = 4,391) | 43.8 (N = 91) | $t_{94.4} = 5.2, P < 0.0001$ |
| Ct value | 26.7 (N = 140) | 24.8 (N = 10) | $t_{10.1} = -0.8, P = 0.45$ |
| Total contacts | 36 (N = 471) | 79 (N = 59) | $\chi^2_1 = 56.6, P < 0.0001$ |

Symptomatic cases had a higher mean number of infected contacts (2.1) compared to asymptomatic cases (0.7) (Table 4). The dispersion parameter *k* was higher for symptomatic cases than asymptomatic cases (0.43 vs. 0.11), indicating lower variance in the number of secondary cases produced by a symptomatic case. This variance is exemplified by the lower percentage of non-transmitters (44%) and higher percentage of superspreaders (9%) among symptomatic cases compared to asymptomatic cases (79% and 4%, respectively). Compared to other age groups, individuals aged 49–64 years had the highest mean number of infected contacts (1.2), lower variance (higher *k*, 0.43), and a higher percentage of superspreaders (3%). Data on total reported contacts showed a different pattern, with a higher mean number of infected contacts (8) as well as higher variance (lower *k*, 0.28) among index cases with >100 total contacts compared to individuals with fewer contacts. This was accompanied by a substantially higher percentage of superspreaders (28%) among individuals with >100 total contacts compared to individuals with 11–100 contacts (19%) or those with 0–10 contacts (2%). Considering only symptomatic adults with a known number of total contacts (N = 129), the percentage of superspreaders was consistently smaller as the number of contacts decreased: 26% (5/19) for individuals with over 100 contacts, 24% (8/34) for those with 21–100 contacts, 8% (2/24) for those with 11–20 contacts, and 0% for those with 10 or fewer contacts (0/52).

**Table 4.** Summary statistics describing the distribution of secondary cases among individual SARS-CoV-2 index cases occurring between December 2019 and July 2021 reported in the literature across 259 studies.

| **Data** | **Sample size** | **Percentage with 0 contacts infected** | **Percentage with 1–5 contacts infected** | **Percentage with >5 contacts infected** | **Maximum contacts infected** | **Estimated mean contacts infected (95% CI)** | **Estimated overdispersion, $k$ (95% CI)** |
| --- | --- | --- | --- | --- | --- | --- | --- |
| All rows | 9,591 | 67% | 30% | 3% | 101 | 0.88 (0.84–0.92) | 0.27 (0.25–0.28) |
| Female | 1,866 | 75% | 22% | 2% | 30 | 0.63 (0.56–0.71) | 0.18 (0.16–0.21) |
| Male | 2,779 | 74% | 24% | 2% | 101 | 0.76 (0.69–0.84) | 0.17 (0.15–0.19) |
| Asymptomatic | 214 | 79% | 17% | 4% | 25 | 0.75 (0.43–1.08) | 0.11 (0.07–0.16) |
| Symptomatic | 697 | 44% | 47% | 9% | 81 | 2.06 (1.8–2.3) | 0.43 (0.36–0.49) |
| Age $\leq 4$ years | 143 | 90% | 10% | 0% | 3 | 0.2 (0.07–0.32) | 0.09 (0.01–0.17) |
| Age 5–12 years | 318 | 97% | 3% | 0% | 3 | 0.04 (0.006–0.07) | 0.03 (–0.006–0.06) |
| Age 13–18 years | 333 | 94% | 6% | 1% | 26 | 0.23 (0.08–0.38) | 0.03 (0.01–0.05) |
| Age 19–24 years | 482 | 88% | 10% | 2% | 19 | 0.38 (0.23–0.52) | 0.06 (0.04–0.09) |
| Age 25–48 years | 2,195 | 76% | 22% | 2% | 101 | 0.73 (0.65–0.82) | 0.15 (0.13–0.17) |
| Age 49–64 | 727 | 54% | 43% | 3% | 35 | 1.16 (1.01–1.31) | 0.43 (0.36–0.51) |
| Age $\geq 65$ years | 284 | 58% | 39% | 3% | 18 | 1 (0.79–1.21) | 0.43 (0.3–0.57) |
| Ct value $\leq 25$ | 67 | 42% | 52% | 6% | 12 | 1.48 (1–1.96) | 0.86 (0.31–1.42) |
| Ct value $> 25$ | 83 | 52% | 41% | 7% | 26 | 1.45 (0.87–2.02) | 0.4 (0.2–0.6) |
| 0–10 total contacts | 281 | 49% | 48% | 2% | 9 | 1.12 (0.93–1.31) | 0.83 (0.53–1.14) |
| 11–100 total contacts | 195 | 47% | 34% | 19% | 39 | 3.1 (2.29–3.91) | 0.32 (0.23–0.41) |
| 101–1000 total contacts | 54 | 35% | 37% | 28% | 101 | 8 (3.92–12.07) | 0.28 (0.16–0.4) |

## DISCUSSION

In this systematic review and meta-analysis, we aimed to characterize the heterogenetity in SARS-CoV-2 transmission among different settings and across individuals that has been reported in published studies. Regarding transmission settings, our meta-analysis identified substantial heterogeneity in attack rates across 12 chosen event types, with higher mean attack rates in nursing homes, cruise ships, households, and other congregate housing settings compared to shopping, hospitals and healthcare, other transportation, and schools. Regarding individual transmission heterogeneity, we found that most cases did not transmit to another person and that a small proportion (3%) of individuals were superspreaders (causing >5 secondary cases). While data on the demographics of index cases were not consistently reported in the literature, the data that were available indicate that superspreaders were more likely to be symptomatic than non-superspreaders, more likely to be adults (with particular overrepresentation in the 49-64 age group), and had more total contacts.

Our ranking of event types by attack rate reinforces our existing understanding of SARS-CoV-2, that transmission is more likely in dense indoor gatherings or close and frequent contact among co-living individuals, especially in households [15]. Published meta-analyses covering the early pandemic (pre-2021) estimated pooled household secondary attack rates of 17–21% [16,18,19,30,31], with household attack rates consistently higher than those in healthcare, work, or travel settings [16,19]. Our pooled household secondary attack rate over 115 events was 29%, higher than these earlier studies but similar to the 31% estimate from Madewell et al. [18] for studies covering July 2020 to March 2021. The higher value may be explained by the emergence of the Alpha and Delta variants and the larger second and third waves of the pandemic occurring in some countries during 2021.

The literature on SARS-CoV-2 transmission events rarely reported on the epidemiological context and characteristics of different populations exposed, which could help explain variation in attack rates. While the timing and location of events may help to explain some of the variation within event types, the remaining variation could depend on event duration (as shown by Supplementary Figure S5) and time spent indoors, types of activities occurring (e.g., exercise, singing) [32,33], and the age groups present at the event. For example, the age of individuals interacting in these contexts appears to also influence propensity for transmission, as evidenced by the large difference in attack rates within schools versus nursing homes. Children and adolescents are frequently found to have lower household infection risk than working age adults [18,19,21,31] and older adults have higher risk of infection and severe disease than younger ages [18,31]. In studies that assessed transmission among school-aged children, teachers, and their household contacts, attack rates among children at school were lower than among teachers and the household contacts of children and teachers [34,35]. Variation in the stringency of interventions (e.g. masking requirements, physical distancing, lockdowns) across countries and over time also could have affected attack rates across different settings. As shown in Supplementary Figure S6 comparing attack rates for events in the United States and China, two locations where differing stringency of control measures were implemented, meta-analysis estimated attack rates were lower across event types for China, though the largest differences between countries were observed for transmission in social venues and mixed settings. Environmental factors such as humidity, room size, ventilation, and air flow [5] could also augment transmission across settings but these were very rarely reported in the literature.

Analysis of index case demographics also highlighted age as an important factor in SARS-CoV-2 transmission and superspreading. While age was only reported in 46% of index cases, nearly all superspreading individuals were adults and there were no reported superspreaders 12 years of age and under, which is consistent with other reviews of SARS-CoV-2 superspreading [36]. Individual and age-related heterogeneity in the amount and assortative patterns of social contacts likely influence superspreading as well. Evidence supports lower transmission from children compared to adults, but effect sizes have been small in some studies [16,21,30,37]. Remaining heterogeneity in individual infectiousness may derive from differences in genetic susceptibility [38,39], body size (accounting for age) [40], baseline lung volume and function [41], immunocompromising disease or co-infection [42,43], or the loudness and wetness of speech [32]. The relative importance of these characteristics to SARS-CoV-2 transmission at a population level are unknown and may be challenging to measure and report at scale. Future work on COVID-19 and other respiratory diseases should address these hypotheses.

Our results indicate substantial heterogeneity in transmission from individuals, observed in other studies [37,41,44], and evidenced by the skewed degree distribution for index cases and the estimate of the dispersion parameter *k*. Our estimate of *k* (0.27, CI: 0.25–0.28) is within the range of previous estimates for a similar period of the pandemic, with values frequently in the range of 0.1–0.7 [3,7,8,27,45]. Caution should be taken when interpreting *k* values, which are sensitive to changes in the tails of a distribution, such as superspreaders or individuals that cause no secondary infections. Without robust isolated case finding and follow-up, contact tracing efforts may undercount the number of zeroes, biasing *k* upwards [46,47]. Alternatively, backwards contact tracing may be susceptible to attachment bias, where infections are preferentially attributed to a known superspreader rather than a separate (known or unknown) transmitter [47]. Additionally, there may be publication bias or more complete contact tracing for large outbreaks with an individual superspreader or with high attack rates [15,47]. These effects would bias *k* downwards and inflate meta-analysis estimated attack rates across event types. It may also lead to the overestimation of the proportion of index cases that are superspreaders.

Without knowledge of the relative impact of these biases, it is challenging to interpret whether *k* is a true representation of SARS-CoV-2 transmission heterogeneity. To improve inference on individual heterogeneity of transmission from outbreak investigations, we recommend that contact tracing efforts use both backward and forward contact tracing [15,21,48], with sufficient follow-up time to identify non-infecting individuals, and complete reporting of contact tracing efforts (e.g., anonymized line lists with infector-infectee and other demographic information).

While our systematic review is the most comprehensive assessment of SARS-CoV-2 superspreading to date, a principal limitation of our analysis was the incomplete data available in the published literature. Beyond information provided about the timing and location of events, very few studies reported any demographics of the exposed individuals, their COVID-19 vaccination status (once introduced) or history of prior SARS-CoV-2 infection, or the density and amount of time indoors. For individual index cases, some studies reported demographic information and the presence/absence of symptoms, but this atypical. We also experienced difficulty with deducing whether contact tracing was performed for all reported cases in transmission chains, especially for terminal nodes. It was not always clear whether cases did not transmit or whether data were missing due to lack of contact tracing, so these cases had to be omitted from the analysis. Testing and tracing policies likely differed between countries, which would affect the collection of index cases that ended up in our review. For this reason, data on index cases are missing from many countries and transmission chains from some countries may be less complete than others. Similarly, the effectiveness of testing and tracing policies varies across settings (e.g., easier in households than large social gatherings), which affects the completeness of transmission chains and likely which outbreaks get published. There were numerous papers that we reviewed with transmission chains that were simply too incomplete or uncertain for us to extract index case data from them. However, without reporting of testing and tracing policies or the effectiveness of tracing efforts within each paper, or a comprehensive database or systematic review of this information in the literature, these remain as uncertainties that must be addressed with better data.

Another limitation of this review was the wide variation in case detection methods across studies. Not all studies reported the total number of contacts that were tested from events and we assumed in the missing cases that the number tested was the same as number exposed. Our sensitivity analysis, using total exposed contacts for all events as the denominator for attack rates instead of total tested contacts, showed that estimated mean attack rates were consistently lower across event types but the ranking of event types was relatively stable (Supplementary Figure S7). However, some studies reported only symptomatic cases or only performed diagnostic tests (e.g., PCR) on symptomatic individuals, thereby missing all reporting of asymptomatic or pausisymptomatic individuals and any secondary cases produced. These missing contacts may be undercounted for both the numerator (contacts that are infected but asymptomatic) and the denominator (including contacts that are asymptomatic and uninfected), which could move attack rates in either direction. Limiting testing to symptomatic contacts has a more predictable effect on individual case degree distributions, reducing the apparent proportion of individuals that transmit and the total secondary cases among individuals that do transmit. Case ascertainment also likely varied by event setting, contributing additional uncertainty in estimated attack rates. For example, performing contact tracing and testing a greater number of contacts was probably easier in settings with consistent or recorded populations like households, schools, and nursing homes than in large social venues like nightclubs. Differences in estimated attack rates by event type may be less drastic than we observed if case ascertainment could be properly addressed with additional ground truth data, i.e., community asymptomatic testing.

Since case detection depends partly on presence of symptoms, some care should be taken in interpreting the finding that superspreaders were more likely to have symptoms than non-superspreaders. We performed an additional analysis on the presence of symptoms across different demographic factors reported in papers (see Supplementary Table S6). The only trend we saw was for age, where the presence of symptoms was somewhat higher for older adults (49 and older). This may have slightly skewed detection of superspreaders among older adults.

However, there were still hundreds of children with symptoms reviewed as index cases, so there were ample opportunities for them to be identified as superspreaders. Therefore, we remain confident in our findings about the rarity of superspreaders among children. However, data from human challenge trials with SARS-CoV-2 have shown that individuals with the highest viral emissions did not have the most severe symptoms, but these super-emitters were also not asymptomatic [41]. These super-emitters, and the majority of superspreaders reported in the literature, tend to have mild to moderate symptoms [36,41]. While the importance of asymptomatic transmission of SARS-CoV-2 should be acknowledged, numerous studies have shown that transmission is more likely from symptomatic individuals compared to completely asymptomatic individuals [18,21,49–51]. However, additional studies that overcome issues of case ascertainment should be done to assess the role of asymptomatic individuals in SARS-CoV-2 superspreading.

To improve the field and our understanding of the drivers of heterogeneity in transmission, we propose standard and consistent reporting on transmission for all outbreaks, as feasible, including details on the epidemiological context of transmission events and complete line lists of cases following contact tracing, with information on case demographics (age, sex, occupation), diagnosis (presence/absence of symptoms, symptom description, test date and results), the duration of contact tracing, and the total number of contacts and the demographic information for contacts (see Appendix 2). Details on the duration of contact tracing should include the entire time period of case finding and how long cases were followed to detect any secondary cases. We recognize the challenge of collecting, storing, and sharing identifiable data from outbreak investigations while continuing to assure confidentiality and improve trust in the health system. However, developing such a reporting system should be a priority for public health as the information has important inplications for reducing the spread of infectious pathogens.

Our comprehensive review found substantial heterogeneity in the transmission of SARS-CoV-2, highlighting the settings and individual characteristics that might be most important to target for controlling superspreading. Secondary attack rates were highest in co-living situations where prolonged contact between individuals facilitated transmission, though there was substantial variation in attack rates within similar settings that remained unexplained and could be disentangled in future meta-analyses focused on the relative influence of built environment, social setting, and control measures on transmission. Given the moderate attack rates among minors in school and the rarity of children among superspreaders, interventions targeting these age groups may be less efficient at preventing SARS-CoV-2 superspreading and could be deprioritized in favor of interventions focusing on adults [21,52], especially those with symptoms and individuals with many daily close contacts. Acknowledging that there remain substantial gaps in data that limit our inference about superspreading, we advocate for consistent reporting on infectious disease outbreaks, ideally with detailed line lists, to facilitate knowledge synthesis about transmission patterns and superspreading in the future. Our review only covered the first phase of the pandemic, so important questions remain about whether patterns in attack rates and individual-level transmission still apply to later pandemic phases with significant population-level immunity. Enhanced reporting of outbreak data would expedite such future investigations.

## DATA AVAILABILITY

All the data were from publicly available databases. The complete database of extracted information from included studies is provided in Appendix 3.

