## Supplementary Material for "Superspreading of SARS-CoV-2: a systematic review and meta-analysis of event attack rates and individual transmission patterns"

### Epidemiology and Infection

*Supplementary Material*

**Removal of duplicate papers**

After importation of papers into EndNote, the name of the searched database (WHO, PubMed, Embase, I Love Evidence) was added as a custom field and results were manually deduplicated. The first comparison was conducted among references with the same title, year, and volume, but different author fields. When author differences were minor (e.g., initials rather than full name) and page numbers matched, we removed one of the duplicates. When one of the matching duplicates was from the WHO COVID database, we removed the other duplicate. If the duplicates were from one of the other three databases, we removed whichever was highlighted by EndNote (i.e., a record with identical metadata) or had less complete metadata. The second comparison was conducted among references with the same author, year, and volume, with different title fields. When the differences in title were minor (e.g., square brackets around title and alpha vs. α) and their page numbers matched, we removed one of the duplicates. Duplicate removal followed the same process as for the first comparison. The third comparison involved manually scanning the list sorted by title. When duplicates were a preprint and a published version, we removed the preprint version unless it was from the WHO database. We checked every preprint to see if it was ultimately published and replaced the preprint if so.

**Screening of papers and data extraction**

Two reviewers independently screened each title and abstract in Covidence (Veritas Health Innovation, Melbourne, Australia), blinded to each other’s choices. When there were discrepancies, we discussed the conflicts until a consensus was reached. We conducted full text review in the same manner as title and abstract screening. We included all papers in languages spoken by our study team, including English, French, German, Mandarin, and Spanish. We included systematic reviews at this phase to ensure that we had captured all relevant papers from those reviews in the data extraction phase. Since multiple papers used the same underlying datasets for their analyses (e.g., the Diamond Princess cruise ship), two reviewers checked for duplication of datasets across papers by geographic locations to ensure that results were only included once in the analysis (Supplementary Table S2).

Each full text article eligible for data extraction was reviewed by one member of the study team; 10% of studies were independently extracted by a second reviewer for quality assurance. Where there was uncertainty in how to extract data, reviewers flagged the article, which was subsequently reviewed by additional study team members and/or discussed at the weekly research group meeting to reach consensus about how data extraction should occur. Where additional information was required to determine appropriate data extraction methods, the manuscript’s corresponding author was contacted by email up to two times.

**Study risk of bias assessment**

The primary risk of bias for our study was inclusion of results where the true attack rates or secondary infections were not reliably ascertained. To mitigate this, we excluded studies of events with a reported testing rate below 50% and identified the events with attack rates above the 75^th^ percentile for each event type for further team review. Similarly, we excluded studies for which authors expressed uncertainty in the reported transmission. We also excluded studies if only individual-level data was present and the chains of infection could not be reliably ascertained, i.e., most cases had more than one possible source of infection. Studies that reported only the prevalence of SARS-CoV-2 infection in a setting without attributing the cases to a setting-specific transmission event (e.g., reporting of teachers and student infected at one time without clarifying in-school versus household transmission) were also excluded as we could not determine an attack rate. We excluded studies for which only the test positivity rate was reported if we could not determine that each individual was only tested once. Studies were also excluded if the majority of individuals were in quarantine or isolation without any contacts, as attack rates and transmission could not be analyzed without exposure. We did not extract studies that had implausible transmission, such as where individuals were considered the source of their own infection.

###### **Sensitivity analyses**

All descriptive analysis and meta-analysis described above were performed using secondary attack rates calculated with the total tested individuals as the denominator. These procedures were repeated using secondary attack rates calculated with the total exposed individuals as the denominator (generally larger than or equal to the total tested) to assess any qualitative differences in the results.

In the review of individual index cases, we noted the presence of “loops” in some transmission trees, wherein there was uncertainty about the identity of the infector of a secondary case and potentially multiple infectors. Index cases included in these loops were removed prior to the analysis of secondary case distributions described above. We assessed the sensitivity of the results to this choice with three alternative treatments of “loops”: 1) retention of index cases involved in loops, 2) removal of all index cases associated with a given paper if any loops were reported, or 3) inclusion of index cases involved in loops as binomial random variables with $p =1-(1/{(x+1))}$, where $x$ is the reported number of secondary cases, and simulating 100 different draws of the secondary cases produced by each “loop” index case. Parameter estimates for the negative binomial distribution were estimated with each of these alternative datasets and all statistical tests (Chi-square, Student’s t, Kruskal-Wallis) described above were repeated.

Meta-analysis results for secondary attack rates were qualitatively similar if the total number of individuals exposed at an event was used as the denominator for calculating secondary attack rates (Supplementary Figures S7 and S8) compared to the results using the total exposed contacts tested as the denominator (Figure 2 and Supplementary Figure S5). The estimated mean attack rates were lower across all categories using the total exposed as the denominator because the latter number was generally larger than the total tested, but the ranking of different categories showed few changes. The top three categories in both models were other congregate housing, household social events, and households; the ranking of nursing homes shifted from the 6^th^ ranked category to the 4^th^ and hospital/healthcare shifted from 11^th^ to 10^th^. Sensitivity analysis on the treatment of “loops” in the description of secondary cases produced by index cases showed that the results were mostly robust to different strategies for dealing with loops. There was some variation in the fitted values of the negative binomial distribution, with the estimated mean varying from 0.87 to 0.94 and the dispersion parameter *k* varying from 0.22 to 0.29. All statistical tests produced results consistent with the original strategy (Supplementary Table S5). Slopes for the relationship between secondary attack rates and event duration across the 12 event types were very similar when using total exposed contacts tested (Supplementary Figure S5) versus total exposed contacts (Supplementary Figure S8) as the denominator, with minor changes. The slope for cruise ships shifted from positive to negative while the slope for other congregate housing shifted from negative to positive.

### Supplementary Table S1. Search strategies for PubMed, WHO COVID database, I Love Evidence COVID database, and Embase databases. Searched on 9 September 2021.

| **Information source** | **Search #** | **Search strategy** | **Results** |
| --- | --- | --- | --- |
| PubMed | #1 | "SARS-CoV-2"[mh] OR "SARS-CoV-2" OR "severe acute respiratory syndrome coronavirus 2" OR "SARSCoV2" OR "novel coronavirus" OR "nCoV" OR "2019 coronavirus" OR "coronavirus 2" OR "COVID-19/transmission"[mh] OR "COVID"[tiab] OR "COVID19" OR "coronavirus disease 2019" | 171463 |
| PubMed | #2 | "superspreader" OR "super spreader" OR "superspreaders" OR "super spreaders" OR "superspreading" OR "super spreading" OR "superemitter" OR "superemitters" OR "disease cluster" OR "disease clusters" OR "disease clustering" OR "infection cluster" OR "infection clusters" OR "infection clustering" OR "cluster analysis"[mh] OR "cluster analysis" OR "disease hotspot"[mh] OR "disease hotspot" OR "Disease Outbreaks/transmission"[mh:noexp] OR "Space-time clustering"[mh:noexp] OR "space-time clustering"[tiab] OR "transmission patterns" OR "transmission dynamics" OR "transmission chain" OR "transmission chains" OR "Contact Tracing/statistics and numerical data"[mh] OR "contact tracing"[tiab] | 95979 |
| PubMed | #3 | ("model"[ti] OR "modeling"[ti] OR "modelling"[ti] OR "models"[ti] OR "Models, Biological"[mh] OR "Models, statistical"[mh] OR "editorial"[pt]) | 2228294 |
| PubMed | #4 | #1 AND #2 NOT #3 | 2233 |
| WHO COVID database | #1 | TW:"superspreader" OR TW:"super spreader" OR TW:"superspreaders" OR TW:"super spreaders" OR TW:"superspreading" OR TW:"super spreading" OR TW:"superemitter" OR TW:"superemitters" OR TW:"disease cluster" OR TW:"disease clusters" OR TW:"disease clustering" OR TW:"infection cluster" OR TW:"infection clusters" OR TW:"infection clustering" OR MH:"cluster analysis" OR TW:"cluster analysis" OR MH:"disease hotspot" OR TW:"disease hotspot" OR MH:"disease outbreaks" OR MH:"space-time clustering" OR TW:"space-time clustering" OR TW:"transmission patterns" OR TW:"transmission dynamics" OR TW:"transmission chain" OR TW:"transmission chains" OR MH:"contact tracing" OR TW:"contact tracing" | 7516 |
| WHO COVID database | #2 | TI:"model" OR TI:"modeling" OR TI:"modelling" OR TI:"models" OR MH:"models, biological" OR MH:"models, statistical" OR PT:"editorial" | 30535 |
| WHO COVID database | #3 | #1 AND NOT #2 | 6248 |
| I Love Evidence COVID database | #1 | "superspreader" OR "super spreader" OR "superspreaders" OR "super spreaders" OR "superspreading" OR "super spreading" OR "superemitter" OR "superemitters" OR "disease cluster" OR "disease clusters" OR "disease clustering" OR "infection cluster" OR "infection clusters" OR "infection clustering" OR "cluster analysis" OR "disease hotspot" OR "Disease Outbreak" OR "Space-time clustering" OR "transmission patterns" OR "transmission dynamics" OR "transmission chain" OR "transmission chains" OR "contact tracing" | 4130 |
| I Love Evidence COVID database | #2 | "model" OR "modeling" OR "modelling" OR "models" OR "editorial" | 32919 |
| I Love Evidence COVID database | #3 | #1 NOT #2 | 2566 |
| Embase | #1 | 'severe acute respiratory syndrome coronavirus 2'/exp OR 'SARS-CoV-2' OR 'severe acute respiratory syndrome coronavirus 2' OR 'SARSCoV2' OR 'novel coronavirus' OR 'nCoV' OR '2019 coronavirus' OR 'coronavirus 2' OR 'coronavirus disease 2019'/exp OR 'coronavirus disease 2019' OR 'COVID':ti,ab OR 'COVID19' | 181621 |
| Embase | #2 | 'superspreader'/exp OR 'superspreader' OR 'super spreader' OR 'superspreaders' OR 'super spreaders' OR 'superspreading event'/exp OR 'superspreading' OR 'super spreading' OR 'superemitter' OR 'superemitters' OR 'disease cluster' OR 'disease clusters' OR 'disease clustering' OR 'infection cluster' OR 'infection clusters' OR 'infection clustering' OR 'cluster analysis'/exp OR 'cluster analysis' OR 'disease hotspot'/exp OR 'disease hotspot' OR 'spatiotemporal analysis'/exp OR 'space-time clustering' OR 'transmission patterns' OR 'transmission dynamics' OR 'transmission chain' OR 'transmission chains' OR 'Contact examination'/exp OR 'contact tracing':ti,ab | 98557 |
| Embase | #3 | 'model':ti OR 'modeling':ti OR 'modelling':ti OR 'models':ti OR 'biological model'/exp OR 'statistical model'/exp OR 'editorial'/exp OR 'conference abstract'/it OR 'editorial'/it OR 'letter'/it OR 'note'/it | 8939677 |
| Embase | #4 | #1 AND #2 NOT #3 | 2585 |

**Supplementary Table S2.** Identification of duplicate datasets that were used in more than one study and determination of overlapping information on transmission.

| **Search word** | **Search in** | **Study** | **Time period*** | **Data contained (transmission chains/attack rate [AR])** | **Data source** | **Exclude** |
| --- | --- | --- | --- | --- | --- | --- |
| Diamond Princess (DP) | Any field | Yang, HY et al. 2020 [1] | 01/20/2020–02/21/2020 | (1) DP, 01/20/2020–02/21/2020: 634 out of the 3711 people on board tested positive. Presented numbers of cases by nationalities, age, and gender, and diagnosis date (Table 1). **Denominator not available for AR.**  (2) A department store in Tianjin, 01/20/2020–02/20/2020: 25 primary cases (6 salespersons + 19 customers). Case #1 was a salesperson, fever on 1/21, confirmed on 2/1. Case #2 contacted Case #1, #3, # 4 when shopping on 1/23. Case #5 was family member of Case #3. **This case is covered in Chen, T et al. 2020 [2].** | Health Commissions, government news, WHO | Yes |
|  |  | Plucinski, MM et al. 2021 [3] | 01/20/2020–02/03/2020 | They “attempted to survey all Americans on the January 20–February 3 sailing of the DP.” 114 out of 437 American passengers and crew tested positive. “The survey response rate was 229/437 (52%).” **ARs by cabin.** | Participants were interviewed February 28–March 18 | No |
|  |  | Mizumoto, K, and G Chowell et al. 2020 [4] | 01/20/2020–02/22/2020 | “A summary of the COVID-19 confirmed cases by age group and symptom status onboard the Princess Cruises Ship is illustrated in Table 2. A total of 531 people had tested positive for the illness as of February 5, 2020. Out of 531 cases, three cases were aged 0–19 years, 117 were aged 20–58 years and 411 were aged 60 years and older.” **Overall ARs. ARs by age.** | [https://www.niid.go.jp/niid/en/2019–ncov–e/9407-covid-dp-fe-01.html](https://www.niid.go.jp/niid/en/2019-ncov-e/9407-covid-dp-fe-01.html) | No |
| Hong Kong | Any field | Cheng, VCC et al. 2020 [5] | 12/31/2019–02/10/2020 | View Hong Kong as an event place. 11 HCWs exposed, 0 infected. | 43 public hospitals | No |
|  |  | Kwok, KO et al. 2020 [6] | 01/23/2020–02/13/2020 | The first 53 cases reported by the Centre for Health Protection (CHP). **Transmission chains among the 53 cases belonging to 10 clusters (Figure 4).** | CHP | No |
|  |  | Lai, CKC et al. 2020 [7] | 01/23/2020–03/01/2020 | The first 100 cases reported by the CHP. **ARs by event:**   1. Lunar-New-Year family dinner party on 1/26 in a seafood restaurant dining hall in North Point: index case (#46) had 18 close contacts and 115 other contacts. Further infected a family member (#42) and a domestic helper (#61) in another household. **Did not match any transmission chains in the 10 clusters from (Kwok, KO et al. 2020).** 2. Lunar-New-Year hot pot party on 1/26 in a party room in Kwun Tong: 19 attendees, 11 family members (#27, 29–37 and 41) were infected. – **Use Lam, HY et al. 2020 [8] for this cluster.** | CHP | No |
|  |  | Adam, DC et al. 2020 [9] | 01/23/2020–05/07/2020 | 1,038 cases reported by the CHP. **Transmission chains by event (Fig. 2):**   1. **‘bar and band’ cluster of undetermined source (n = 106).** 2. **A wedding** 3. **A temple cluster** 4. **Other clusters where the source and transmission chain could be determined** | CHP | No |
|  |  | Wong, NS et al. 2020 [10] | 01/23/2020–06/19/2020 | 1,128 cases reported by the CHP. **Insufficient information to determine “who infected whom” from Figure 2. We contacted the corresponding author and received the response "The identity and direction of infections are not known. And we are not able to share the original data as they belong to the Hospital Authority and the Department of Health, HKSAR Government."** | CHP | Yes |
|  |  | Lam, HY et al. 2020 [8] | 01/26/2020–02/29/2020 | A Chinese New Year gathering on 1/26 in a commercial party room: 19 attendees, 12 confirmed cases. The attendees had an indoor hotpot dinner and a barbeque held at an outdoor area. **Reported age and sex of each case.** | CHP | No |
|  |  | Wong, SCY et al. 2020 [11] | 02/01/2020–02/29/2020 | An outbreak in Ward A in the Accident and Emergency Service Department at Queen Elizabeth Hospital. The index patient: a 64-year-old woman. All specimens from the 52 contacts were negative. | Queen Elizabeth Hospital | No |
|  |  | Chu, DKW et al. 2021 [12] | March 2021 | A superspreading event at a fitness center. | a local health authority | No |
| Tianjin | Any field | Zhang, Y et al. 2020a [13] | 01/11/2020–01/29/2020 | 17 cases in a workplace. Index case: male, 58 years old, developed fever on 1/14, confirmed on 01/21/2020. | Tianjin CDC | No |
|  |  | Wang, J et al. 2020 [14] | 01/14/2020–02/20/2020 | Transmission chains for 131 cases (FIGURE 3) including two clusters:   1. **Two train conductors infected 17 people** 2. **A shopping mall in Baodi District**    a. 45 total cases    b. Index case: a 35-years-old saleswoman (#43), fever on 1/21    c. Infected one family member, 5 salespersons, and 22 customers. | Municipal People’s Government (<http://www.tj.gov.cn/>), the Tianjin Health Committee | No |
|  |  | Chen, T et al. 2020 [2] | 01/14/2020–03/13/2020 (China Standard Time) | Transmission chains (Fig 2.) of Baodi department store:   a. 43 total cases b. Index case: a saleswoman, fever on 1/21 c. Infected 6 other saleswomen.   **Use Wang, J et al. 2020 [14] for Baodi department store outbreak with more complete data.** | Municipal Health Commission (<http://wsjk.tj.gov.cn/>) | Yes |
|  |  | Liu, YF et al. 2020 [15] | 01/20/2020–02/22/2020 | 115 cases in 33 clusters out of 135 total cases.   1. **A family cluster in Baodi district (Fig 1)**    a. Index case: female, 50 years old, symptom on 2/3, confirmed on 2/10    b. Infected 7 family members 2. **Workplace outbreak (Fig 2)**    a. 10 cases in total, all male, 37–58 years old    b. One index case infected 6 coworkers 3. Department store - **Use Wang, J et al. 2020 [14] for Baodi department store outbreak with more complete data.**    a. 26 cases in total    b. Index case: symptom on 1/21, salesperson    c. Infected 5 salespersons, 20 customers | Chinese CDC | No |
|  |  | Tindale, LC et al. 2020 [16] | 01/20/2020–02/19/2020 | Figure 6, 7 had infector-infectees pairs for Singapore during 1/20/2020–2/16/2020 and for 135 cases in Tianjin during 1/14/2020–2/19/2020. **Extract data from Fig 6 and Fig 7; for case 77, put 0 for infected contacts. Easiest one to extract data for Tianjin’s 135 cases.** | Health Commission of Tianjin; Ministry of Health Singapore | No |
|  |  | Luo, C et al. 2021 [17] | 01/21/2020–02/22/2020 | 135 cases in Tianjin and 143 cases in Chengdu.   1. Figure 1. Transmission network graph for confirmed COVID-19 cases in Tianjin. **- Use Tindale, LC et al. 2020 [16] for Tianjin’s 135 cases.** 2. **Fig 2. Transmission network graph for confirmed COVID-19 cases in Chengdu. *The central node is not a case.** | Municipal Health Commission | No |
|  |  | Li, Y et al. 2021 [18] | 01/21/2020–02/26/2020 | Figure 1. Transmission chains of 135 cases in Tianjin and **942 cases in Hebei Province**. **-Use Tindale, LC et al. 2020 [16] for Tianjin’s 135 cases. Extract Hebei’s data only.** | Health Commission of Tianjin; Health Commission of Hebei | No |
|  |  | Zhang, Y et al. 2020b [19] | 01/21/2020–02/26/2020 | Figure 2. Transmission chains of 135 cases. **-Use Tindale, LC et al. 2020 [16] for Tianjin’s 135 cases.** | Municipal Health Commission (<http://www.tj.gov.cn/>) | Yes |
| Cruzado | Author | Cruzado, MAAS et al. 2020a [20] | 03/14/2020–05/14/2020 | The dialysis unit of a tertiary hospital in the Philippines: 8 out of 167 patients were positive. Published in: Nephrology 2020 Vol. 25. Has transmission chain. | A tertiary hospital in the Philippines | No |
|  |  | Cruzado, MAAS et al. 2020b [21] | 03/14/2020–05/14/2020 | The dialysis unit of a tertiary hospital in the Philippines: 8 out of 167 patients were positive. Published in: Journal of the American Society of Nephrology 2020 Vol. 31. **Unable to find full text.** | A tertiary hospital in the Philippines | Yes |
| Shenzhen | Any field | Bi, Q et al. 2020 [22] | 01/14/2020–02/12/2020 | **Count this as an event. Extract data from Table 3. *The first column has a typo – Number of cases should be Number of contacts.** | Shenzhen Center for Disease Control and Prevention | No |
|  |  | Xu, S et al. 2021 [23] | 01/14/2020–02/29/2020 | Figure 2. Cluster events of COVID-19 in Shenzhen City, 29 February 2020 (n=97). | China Information System for Disease Control and Prevention | No |
|  |  | Huang, F et al. 2021 [24] | 01/19/2020–02/21/2020 | Exclude b/c no available data. | Shenzhen Municipal Health Commission and a COVID-19 diagnosis and treatment fixed-point hospital | Yes |
| Singapore | Any field | Venkatachalam, I et al. 2021 [25] | 01/06/2020–03/16/2020 | 17 out of 543 suspect patients were positive. Denominator is suspect patients not # of exposed. <10% exposed healthcare workers were tested. | Singapore General Hospital (a tertiary hospital) | Yes |
|  |  | Wee, LEI et al. 2020 [26] | 02/07/2020– 05/07/2020 | 28 out of 4,621 patients in the respiratory surveillance ward were positive; 845 out of 2,681 patients in the isolation ward were positive. **Table 1 has number of contacts (total and infected) for each index case. All the index cases were patients.** | Singapore General Hospital (a tertiary hospital) | No |
|  |  | Wee, LE et al. 2020 [27] | 01/01/2020–04/22/2020 | **An outbreak among healthcare workers.** | Singapore General Hospital (a tertiary hospital) | No |
|  |  | Yong, SEF et al. 2020 [28] | 01/19/2020–02/17/2020 | 28 cases from 3 clusters from:   1. Church A: service on **Jan 19**, 2020; index cases were 56-yr-old female (onset: 01-22-2020) and 56-yr-old male (onset: 01-24-2020); infected 4 people. 2. Family gathering on Jan 25, 2020 3. Church B: detected in mid-February   **Figure 1 has transmission chains and Figure 2 has demographics.** | The Ministry of Health | No |
|  |  | Ng, OT et al. 2021 [29] | 01/23/2020–04/03/2020 | 188 out of 7770 close contacts linked to 1114 index cases were positive. **Count this as an event, like Bi, Q et al. 2020 [22]. We can calculate ARs by household, work, and social.** | The Ministry of Health | No |
|  |  | Wei, WE et al. 2020 [30] | 01/23/2020–03/16/2020 | 243 cases; FIGURE has “who infected whom” for 7 clusters:   A. Church cluster: arrived in Singapore on Jan 19. They visited a local church the same day and had symptom onset on Jan 22 (patient A1) and Jan 24 (patient A2). -**Use** Yong, SEF et al. 2020 [28] **with more data for this cluster.** B. Singing class cluster C. Wife infected husband D. Husband infected wife E. Man infected housemate F. Church service on March 1 G. A man traveled to Indonesia Mar 3–7, infected a woman | The Ministry of Health | No |
|  |  | Chen, J et al. 2020 [31] | 01/24/2020–02/15/2020 | 16 out of 335 passengers on a flight from Singapore to Hangzhou were positive. | Interviews of passengers | No |
|  |  | Pung, R et al. 2020 [32] | 02/03/2020–02/15/2020 | 36 cases from 3 clusters:   A. **A tour group** visited a jewelry shop and a complementary health products shop B. **A company conference and subsequent family transmissions**  - 42 years, male, Malaysian (B1), traveled to Singapore on Jan 16–23, 2020, for a company conference - The conference was attended by at least 111 participants from 19 different countries on Jan 20–22, 2020 - 17 attendees at the conference were from mainland China. - B1 infected sister and mother-in-law  C. Visit to church on **Jan 19**: 56 years, male, husband of C2 traveller from Wuhan and 56 years, female, wife of C1 traveller from Wuhan infected 3 other people. -**Use (Yong, SEF et al. 2020) with more data for this cluster.**   **Figure 1 has “who infected whom”.** | The Ministry of Health; “Open source reports were obtained for overseas cases” | No |
|  |  | Pang, J et al. 2020 [33] | 03/21/2020–04/24/2020 | The first nursing home COVID-19 outbreak in Singapore and Figure 4 has household transmission that led to the nursing home outbreak. | Serial point prevalence testing, active contact tracing, screening of close contacts, whole genome sequencing and phylogenetic analysis. | No |
|  |  | Van Gunten, T 2021 [34] | 03/25/2020–04/19/2020 | Cluster 1: a major construction site  Clusters 2 and 3: both dormitories  **We contacted the corresponding author and decided that there was insufficient information to determine “who infected whom” from Figure I.** | The Ministry of Health | Yes |
|  |  | Chew, MH et al. 2020 [35] | 04/11/2020–04/19/2020 | A privately owned dormitory: 1264 out of 1832 symptomatic foreign workers were positive. | Review of investigation results | No |
|  |  | Pung, R et al. 2021 [36] | 01/23/2020–03/21/2020 | “Of the 400 cases identified before 21 March 2020, 46 cases and their households were omitted due to common exposure between cases and household contacts or cases reported to self-isolate from their household contacts… 34 had no household contacts, 277 were primary or co-primary cases in their household and 43 household contacts were tested positive for SARS-CoV-2. A total of … 875 household contacts were identified” *Singapore: the first imported case on 23 January 2020 | Contact tracing | No |
| Malaysia | Any field | Ahmad, NA et al. 2020 [37] | 01/16/2020–02/22/2020 | The first local transmission cluster of COVID-19 in Malaysia:   - Index case: a Malaysian man aged 41-year-old - On Jan 16, 2020, he travelled alone to Singapore - Attended a conference during Jan 20-22. - 59 contacts traced from the index case: two positive cases detected.   **Use Pung, R et al. 2020 [32] for more detailed data.** | National and State Crisis Preparedness and Response Centres, District Health Office | Yes |
|  |  | Chaw, L et al. 2020a [38] | 03/05/2020–04/02/2020 | 19 out of 75 individuals in Brunei who attended the Tablighi event in Malaysia were positive. **Preprint duplicate of “Analysis of SARS-CoV-2 Transmission in Different Settings, Brunei”** | Brunei’s Ministry of Health | Yes |
|  |  | Chaw, L et al. 2020b [39] | 03/05/2020–04/02/2020 | 19 out of 75 individuals in Brunei who attended the Tablighi event in Malaysia were positive. **Figure 2 has “who infected whom”.** | Brunei’s Ministry of Health | No |
| Wuhan | Title | Li, Q et al. 2020 [40] | 12/2019–01/2020 | The first 425 cases in Wuhan. 5 clusters of cases:   1. M61 index (onset: 12/20/2019) infected F57 and F31 2. F62 index (onset: 12/27/2019) infected M64 3. M49 index (onset: 12/12/2019) infected F48, M78, M50 4. F52 index (onset: 12/21/2019) infected M51, F25 5. M32 index (onset: 1/4/2020) infected F28, M57, F? | A joint field epidemiology team | No |
|  |  | Li, F et al. 2021 [41] | 12/2019–04/2020 | 29 578 primary cases and 57 581 household contacts  **Table 2 has ARs among households with a single primary case by sex and age of contacts** | Wuhan Center for Disease Control and Prevention | No |
|  |  | Wang, X et al. 2020 [42] | 01/05/2020–02/12/2020 | **Figure 2 has transmission chains for 9 clusters in the Department of Neurosurgery. Include “probable transmission”.** | Wuhan Union Hospital | No |
|  |  | Zhao, D et al. 2020 [43] | 01/14/2020–02/21/2020 | Individual spreader:   - The index patient was a 59-year-old male, visited wife in a cancer ward. - On 01/03/2020 had fever, headache, chest pain, and myalgias - Died on 26 January - 4 female patients in the same ward as the wife were infected   Event:   - From 01/14 to 02/21, 1407 HCWs were screened - 347 underwent chest CT only, no PCR: unclear how many had abnormal rest among them - 1060 had PCR: 160 were positive | Renmin Hospital of Wuhan University | No |
|  |  | Yi, B et al. 2021 [44] | 01/22/2020–02/04/2020 | **Count all familiar clusters in Wuhan as an event. Extract data from Table 1.** **Has number of index cases + number of infected contacts, but no further information. Insufficient data.** | Renmin Hospital of Wuhan University | Yes |
|  |  | Luo, Y et al. 2020 [45] | 01/31/2020–02/21/2020 | The index patient was a 39-year-old nephrologist at Central Hospital of Wuhan | Zhongnan Hospital of Wuhan University | No |
|  |  | Horchinbilig, U et al. 2020 [46] | 02/17/2020–05/08/2020 | Randomly enrolled 100 cases and the family members they had contact with. “100 families were analyzed…61 patients were identified as index cases, and 83 were diagnosed with SARS-CoV-2 infection contracted from the index cases. **Has number of index cases + number of infected contacts, but no further information. Insufficient data.** | Zhuankou Fangcang Shelter Hospital | Yes |
|  |  | Chen, S et al. 2020 [47] | 01/25/2020–02/27/2020 | 3 cases of a familial cluster with one family member being a kidney transplant recipient | Tongji Hospital | No |
|  |  | Tan, F et al. 2020 [48] | 03/20/2020–04/05/2020 | “After performing contact tracing for all patients, only one patient was found to have transmitted the virus; he passed the virus to his mother... Persons with whom the other 11 patients came into contact were not infected” | People’s Hospital of Wuhan University. | No |
| South Korea | N/A | Park, Y et al. 2020 [49] | 01/20/2020– 03/27/2020 | “We analyzed reports for 59,073 contacts of 5,706 coronavirus disease (COVID-19) index patients reported in South Korea during January 20–March 27, 2020.”  Event data  **Study identifies transmission occurring in South Korea during the period of the first cases in South Korea, using the same data source as Jeong, EK et al. 2020 [50]** | Korea Centers for Disease Control and Prevention (KCDC) | Include event data |
|  |  | Jeong, EK et al. 2020 [50] | 01/24/2020– 03/10/2020 | “Between January 24th and March 10th, a total of 2,370 individuals had contact with the first 30 cases of COVID-19.”  Individual spreader data  **Study identifies the contacts of the first 30 cases in South Korea, using the same data source as Park, Y et al. 2020 [49]** | Korea Centers for Disease Control and Prevention (KCDC) | Include individual spreader data |
| Asan Medical Center | Any field | Jung, J et al. 2021a [51] | 01/01/2020–12/20/2020 | “During the study period, there were 21 nosocomial events at our hospital, which involved 65 individuals with COVID-19 (caregivers, N = 21 (32%); patients, N = 18 (28%); HCWs, N = 17 (26%); family members infected in the community setting, N = 9 (14%)).”  Individual spreader data, 65 with occupation  **Study takes place in Asan Medical Center. The study identifies events that take place during the study period, which overlaps with Jung, J et al. 2021b [52] by roughly 10 months.** | Asan Medical Center | Include |
|  |  | Jung, J et al. 2021b [52] | 03/01/2020–03/01/2021 | “During the study period, there were 440 close contacts and 2,198 non-close contacts from 14 index cases (9 patients or caregivers and 5 healthcare workers [HCWs]...). There were 26 (5.9%) secondary cases from close contacts and 10 (0.5%) from non-close contacts”  More solid transmission tree, symptoms, no dates  **Study takes place in an unnamed tertiary care center that was identified to be Asan Medical Center. The study identifies transmission that take place during the study period, which overlaps with Jung, J et al. 2021a [51] by roughly 10 months.** | Asan Medical Center | Exclude |
| Karnataka | Any Field | Kumar, N et al. 2020 [53] | 03/08/2020– 05/31/2020 | “The study included all cases diagnosed with COVID-19 in the Karnataka state, reported from March 8 – May 31, 2020. Case identification was carried out by the staff of each district, which included the district surveillance officer and their teams, the Integrated Disease Surveillance Programme (IDSP) team, and the urban health authorities of the major cities Table 2 presents the clinical status and outcome of the COVID-19 cases in Karnataka. Overall, amongst the 3404 cases, the final outcome was available for 1394 (41%) at the end of the study period (May 31, 2020)”  **Uses line-list of cases in Karnataka. Presents transmission trees for cases in Karnataka March-May.**  (Verified the case #s and # infected that was previously extracted by Sophie as repeats of Nagarajan, K et al. 2020 [54] data) | Government of Karnataka Health and Family Welfare Services Department, ICMR Portal & Integrated Disease Surveillance Programme (IDSP) | Exclude |
|  |  | Pattabiraman, C et al. 2020 [55] | 03/05/2020–05/21/2020 | “The line list of positive patients was provided by the Directorate of Health and Family Welfare Services, Karnataka and missing data was filled in from the ICMR portal. […] The epidemiological data was extracted from the line list and a contact map was constructed using the state line list of positive cases. […] Karnataka recorded 1578 cases between March 5–May 21, 2020”  **Uses line-list of cases in Karnataka. Presents transmission trees for cases in Karnataka March–May.**  (Verified the case #s and #s infected overlap with Nagarajan, K et al. 2020 [54]) | Government of Karnataka Health and Family Welfare Services Department, ICMR Portal | Exclude |
|  |  | Gupta, M et al. 2020 [56] | 03/09/2020–07/21/2020 & 03/09/2020–06/01/2020** | “We used data generated through surveillance activities undertaken by the Integrated Disease Surveillance Program (IDSP) and the Department of Health and Family Welfare in accordance with national and state policies. Data was de-identified before extraction and analysis. "The first dataset was sourced from daily COVID-19 bulletins released by the Government of Karnataka [...] The second dataset comprised of linelist contact tracing data maintained by the IDSP ". It appears there was an overlap in the merged sources, so the same individual may have appeared in both. From 9 March to 21 July 2020, Karnataka reported 71068 cases”  **Uses line-list of cases in Karnataka. Presents transmission trees for the largest three clusters in Karnataka March–July.** | Government of Karnataka Health and Family Welfare Services Department daily bulletins & Integrated Disease Surveillance Program (IDSP) | Partial include (Bellary cluster) |
|  |  | Saraswathi, S et al. 2020 [57] | 03/09/2020–05/17/2020 | “Daily consolidated bulletins, containing anonymised patient and contact data, were uploaded by the government to the portal it created to share information on COVID-19. […] As of 17 May 2020, Karnataka had declared 1147 diagnosed cases […] For our analysis, we downloaded the daily bulletins containing information for all cases reported positive for COVID-19 from 9 March to 17 May 2020”  **Uses line-list of cases in Karnataka. Presents transmission trees for cases in Karnataka March–May.** | Government of Karnataka Health and Family Welfare Services Department daily bulletins | Exclude |
|  |  | Nagarajan, K et al. 2020 [54] | 03/09/2020–05/23/2020 | “This publicly available contact tracing data of 1959 diagnosed SARS-CoV-2 patients between March 09, 2020, to May 23, 2020, has been collected, compiled and updated by the Department of Health and Family Welfare Services, Government of Karnataka, and other stakeholders for the benefit of the larger public”  **Uses line-list of cases in Karnataka. Presents a transmission tree for cases in Karnataka March–April.**  Links to a line-list with 70k+ cases. Taking data from the csv file up until the end of May, which is when contact tracing starts to get spottier. (March–May is ~3000 cases, March–Jul is ~70,000) | Government of Karnataka Health and Family Welfare Services Department daily bulletins | Partial Include |

*Start date and end date (MM/DD/YYYY) of the data covered in the study. Start date was the earliest date when the event/exposure happened. End date was the latest date they stopped following the contacts. **Study used two data sources with separate end dates.

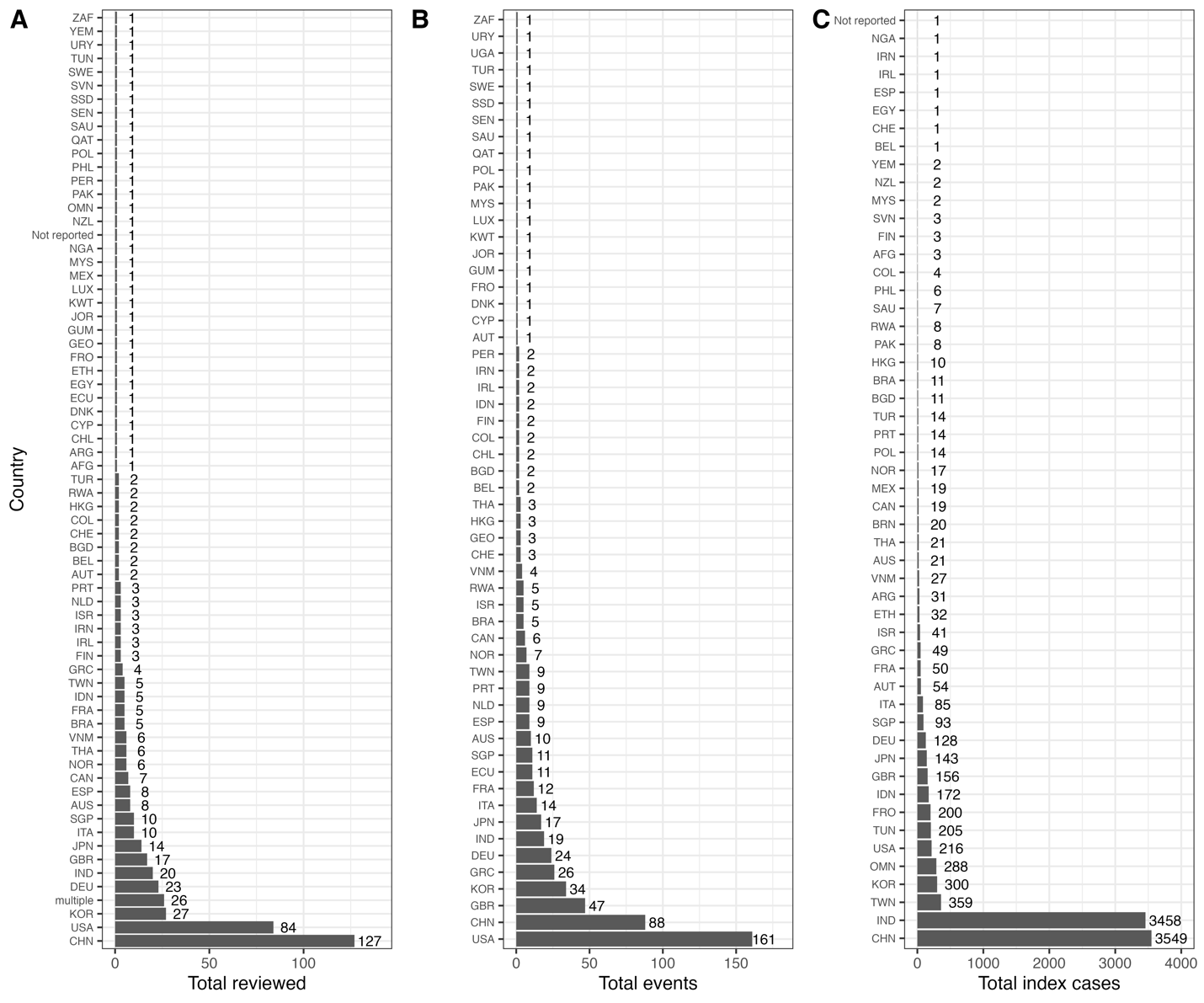

**Supplementary Figure S1.** Focal countries providing data on SARS-CoV-2 transmission across all reviewed papers (A), superspreading events (B), and individual superspreading index cases (C) from December 2019 to August 2021 compiled from literature. Countries are abbreviated using three-letter ISO 3166-1 country codes.

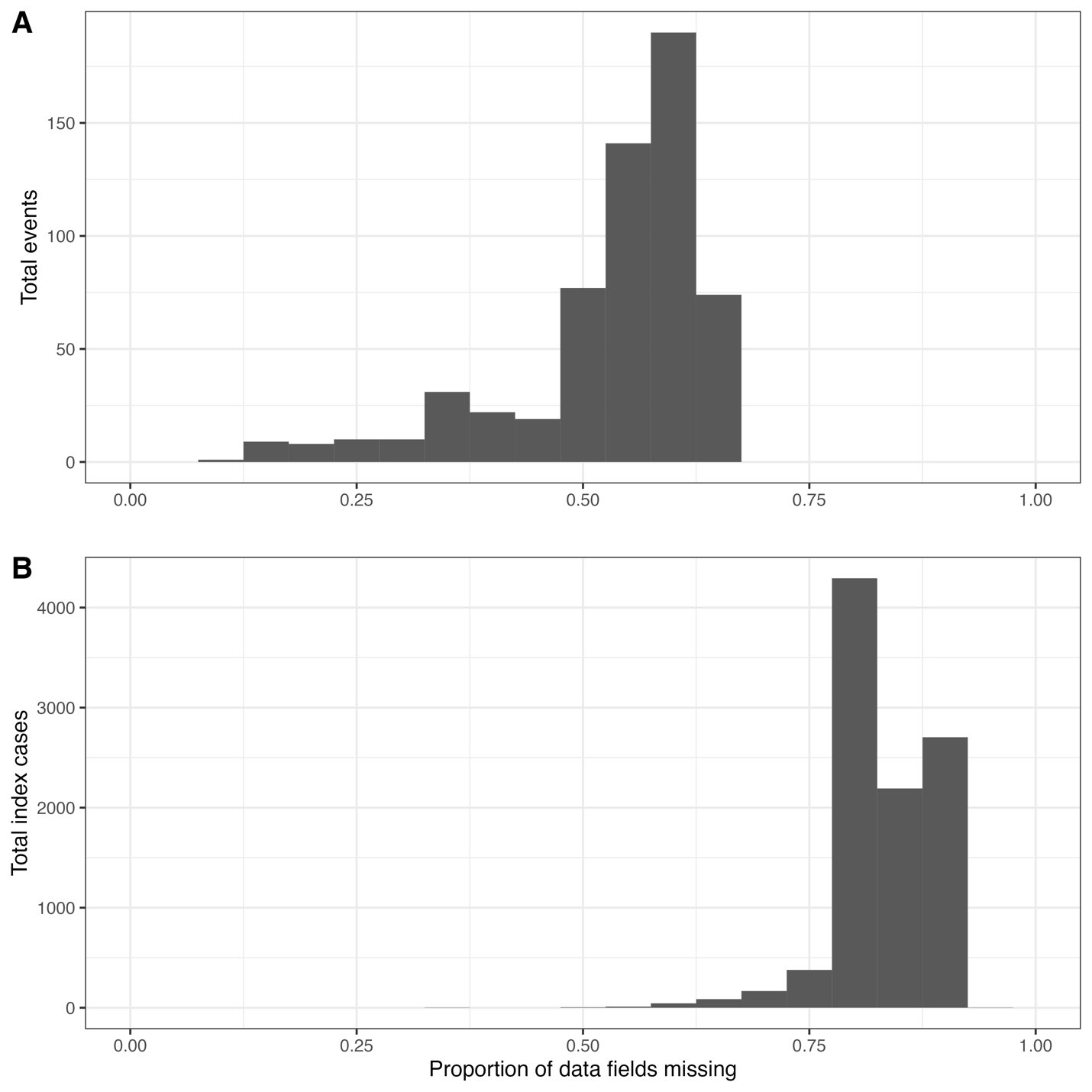

**Supplementary Figure S2.** Missingness of data fields across SARS-CoV-2 superspreading events (A) and individual SARS-CoV-2 index cases (B) occurring between December 2019 to August 2021. Proportions for event data were calculated as the number of data fields with missing information out of the total (N = 46) across all 592 events. Proportions for index cases were calculated as the number of data fields with missing information out of the total (N = 74) across all 9,883 events.

**Supplementary Table S3.** Data dictionary for Google Sheet for recording SARS-CoV-2 superspreading event data for events occurring between December 2019 to August 2021, including the completeness of data reporting for each data field. Completion rates were calculated as the total number of events with a completed data field divided by the total number of events (N = 592).

| **Variable** | **Type** | **Description** | **Number of events with missing data** | **Completion rate (%)** |
| --- | --- | --- | --- | --- |
| paper_id | character | Unique paper ID from Covidence. | 0 | 100 |
| event_id | numeric | Event ID. Start from 1. | 0 | 100 |
| event_type | character | Type of event. An event is defined as emergence of cases within a short time period within a defined geographic location. For example, an outbreak in a hospital ward or a two-day conference in Singapore. | 0 | 100 |
| event_desc | character | Brief description of the event. | 0 | 100 |
| event_indoor | character | The event happened completely indoor, completely outdoor, or both. | 0 | 100 |
| event_start_month | numeric | Start date of the event/exposure - month. | 1 | 100 |
| event_start_day | numeric | Start date of the event/exposure - day. | 1 | 100 |
| event_start_year | numeric | Start date of the event/exposure - year. | 0 | 100 |
| event_end_month | numeric | End date of the event/exposure - month. | 1 | 100 |
| event_end_day | numeric | End date of the event/exposure - day. | 1 | 100 |
| event_end_year | numeric | End date of the event/exposure - year. | 0 | 100 |
| event_country | character | Where did the event happen - country. Select ISO 3166-1 alpha-3 codes from the dropdown list. | 0 | 100 |
| event_admin1 | character | Where did the event happen - 1st level administrative unit/code. | 90 | 85 |
| event_admin2 | character | Where did the event happen - 2nd level administrative unit/code. | 303 | 49 |
| event_admin3 | character | Where did the event happen - 3rd level administrative unit/code (if reported). | 483 | 18 |
| exposed_num | numeric | Total number of exposed people in this event, including primary cases. | 3 | 99 |
| exposed_tested_num | numeric | Total number of exposed people in this event who were tested. | 210 | 65 |
| secondary_cases_num | numeric | Total number of infected at the event. | 0 | 100 |
| secondary_cases_test_method | character | Testing method for the secondary cases. | 43 | 93 |
| secondary_cases_test_method_other | character | Free text. | 499 | 16 |
| exposed_vaccinated | numeric | Among the exposed: number of fully vaccinated people. | 575 | 3 |
| secondary_cases_vaccinated | numeric | Infected cases at the event: number of vaccinated. | 579 | 2 |
| age_grp1 | character | Definition of age group 1, e.g, <10 years | 477 | 19 |
| age_grp1_exposed | numeric | Number of exposed who were within age group 1. | 513 | 13 |
| age_grp1_tested | numeric | Number of exposed who were within age group 1 and were tested. | 549 | 7 |
| age_grp1_infected | numeric | Number of infected who were within age group 1. | 481 | 19 |
| age_grp2 | character | Definition of age group 2, e.g, 10–24 years old | 495 | 16 |
| age_grp2_exposed | numeric | Number of exposed who were within age group 2 | 529 | 11 |
| age_grp2_tested | numeric | Number of exposed who were within age group 2 and were tested. | 559 | 6 |
| age_grp2_infected | numeric | Number of infected who were within age group 2 | 497 | 16 |
| age_grp3 | character | Definition of age group 3, e.g, 25–64 yrs | 527 | 11 |
| age_grp3_exposed | numeric | Number of exposed who were within age group 3 | 556 | 6 |
| age_grp3_tested | numeric | Number of exposed who were within age group 3 and were tested. | 570 | 4 |
| age_grp3_infected | numeric | Number of infected who were within age group 3 | 530 | 10 |
| age_grp4 | character | Definition of age group 4, e.g, >= 65 years old. | 539 | 9 |
| age_grp4_exposed | numeric | Number of exposed who were within age group 4 | 563 | 5 |
| age_grp4_tested | numeric | Number of exposed who were within age group 4 and were tested. | 575 | 3 |
| age_grp4_infected | numeric | Number of infected who were within age group 4 | 542 | 8 |
| exposed_males | numeric | Among the exposed: number of males. | 499 | 16 |
| exposed_males_tested | numeric | Among the exposed: number of males tested. | 499 | 16 |
| secondary_cases_male | numeric | Infected cases at the event: number of males. | 432 | 27 |
| exposed_females | numeric | Among the exposed: number of females. | 509 | 14 |
| exposed_females_tested | numeric | Among the exposed: number of females tested. | 514 | 13 |
| secondary_cases_female | numeric | Infected cases at the event: number of females. | 453 | 23 |
| variant | character | Variant name. | 0 | 100 |
| prevention_context | character | Prevention context. | 259 | 56 |

**Supplementary Table S4.** Data dictionary for Google Sheet for recording individual SARS-CoV-2 index case data from December 2019 to July 2021, including the completeness of data reporting for each data field. Completion rates were calculated as the total number of events with a completed data field divided by the total number of index cases (N = 9,883).

| **Variable** | **Type** | **Description** | **Number of index cases with missing data** | **Completion rate (%)** |
| --- | --- | --- | --- | --- |
| paper_id | character | Unique paper id from Covidence. | 0 | 100 |
| index_case_id | character | Individual spreader/index case ID. For transmission tree, each infected contact becomes an index case. | 0 | 100 |
| index_case_country | character | Index case: Where was the index case when the transmission happened - country. | 0 | 100 |
| index_case_admin1 | character | Index case: Where was the index case when the transmission happened - 1st level administrative unit/code. | 1349 | 86 |
| index_case_admin2 | character | Index case: Where was the index case when the transmission happened - 2nd level administrative unit/code. | 4617 | 53 |
| index_case_admin3 | character | Index case: Where was the index case when the transmission happened - 3rd level administrative unit/code. | 9406 | 5 |
| index_case_onset_date | date | Index case: Date of symptom(s) onset. | 8111 | 18 |
| index_case_symptoms | character | Index case: Symptoms experienced. | 8863 | 10 |
| index_case_pos_test_date | date | Index case: Date of positive test by PCR or other testing method. | 5574 | 44 |
| index_case_test_method | character | Testing method for the index case. | 3290 | 67 |
| index_case_test_method_other | character | Free text. | 9334 | 6 |
| index_case_age_yrs | numeric | Index case: Age (in years). | 5311 | 46 |
| index_case_gender | character | Index case: Gender. | 5095 | 48 |
| index_case_occupation | character | Index case: Occupation. | 9488 | 4 |
| index_case_outcome | character | Index case: Clinical outcome. | 6528 | 34 |
| viral_shedding_ct_value | numeric | Index case: Cycle threshold (CT) value. List lowest value if multiple reported. | 9694 | 2 |
| viral_shedding_specimen_type | character | Index case: Specimen type. | 9704 | 2 |
| contact_definition | character | How was a contact defined. | 7810 | 21 |
| contact_definition_distance | character | Does the contact definition include physical distance? | 9106 | 8 |
| contact_definition_time | character | Does the contact definition include time? | 9049 | 8 |
| contacts_total | numeric | Contacts: Total number of contacts, regardless of type. | 9337 | 6 |
| contacts_total_tested | numeric | Contacts: Total number of contacts, regardless of type, who were tested. | 9570 | 3 |
| contacts_total_infected | numeric | Contacts: Number of infected contacts, regardless of type. | 0 | 100 |
| loop | character | If the index case is part of an infection loop (shares an infectee with another possible infector) | 9591 | 3 |
| contacts_test_method | character | Testing method for the contacts. | 3546 | 64 |
| contacts_test_method_other | character | Free text. | 9378 | 5 |
| contacts_hh | numeric | Contacts: Number of household contacts, defined as those who live together full-time. | 9691 | 2 |
| contacts_hh_tested | numeric | Number of household contacts tested. | 9740 | 1 |
| contacts_hh_infected | numeric | Contacts: Number of infected household contacts. | 8933 | 10 |
| contacts_community | numeric | Contacts: Number of community contacts, defined as any contact that is not household contact. | 9845 | 0 |
| contacts_community_tested | numeric | Number of community contacts tested. | 9868 | 0 |
| contacts_community_infected | numeric | Contacts: Number of infected community contacts, defined as any contact that is not household contact. | 9655 | 2 |
| contacts_other | character | Define what other type of close contacts it is. | 9304 | 6 |
| contacts_other_specify | character | Free text. | 9676 | 2 |
| contacts_other_total | numeric | Other contacts: Total number. | 9736 | 1 |
| contacts_other_tested | numeric | Other contacts: Total number tested. | 9810 | 1 |
| contacts_other_infected | numeric | Other contacts: Total number of infected. | 9364 | 5 |
| contacts_other2 | character | Define what other type of close contacts it is. | 9728 | 2 |
| contacts_other2_specify | character | Free text. | 9840 | 0 |
| contacts_other2_total | numeric | Other contacts: Total number. | 9824 | 1 |
| contacts_other2_tested | numeric | Other contacts: Total number tested. | 9863 | 0 |
| contacts_other2_infected | numeric | Other contacts: Total number of infected. | 9731 | 2 |
| contacts_other3 | character | Define what other type of close contacts it is. | 9875 | 0 |
| contacts_other3_specify | character | Free text. | 9870 | 0 |
| contacts_other3_total | numeric | Other contacts: Total number. | 9879 | 0 |
| contacts_other3_tested | numeric | Other contacts: Total number tested. | 9879 | 0 |
| contacts_other3_infected | numeric | Other contacts: Total number of infected. | 9868 | 0 |
| contacts_other4 | character | Define what other type of close contacts it is. | 9880 | 0 |
| contacts_other4_specify | character | Free text. | 9881 | 0 |
| contacts_other4_total | numeric | Other contacts: Total number. | 9880 | 0 |
| contacts_other4_tested | numeric | Other contacts: Total number tested. | 9880 | 0 |
| contacts_other4_infected | numeric | Other contacts: Total number of infected. | 9880 | 0 |
| contacts_age_grp1 | character | Contacts: Definition of age group 1, e.g, <10 years | 9294 | 6 |
| contacts_age_grp1_total | numeric | Contacts: Number of contacts who were within age group 1. | 9819 | 1 |
| contacts_age_grp1_infected | numeric | Contacts: Number of infected contacts who were within age group 1. | 9304 | 6 |
| contacts_age_grp2 | character | Contacts: Definition of age group 2, e.g, 10–24 years old | 9655 | 2 |
| contacts_age_grp2_total | numeric | Contacts: Number of contacts who were within age group 2 | 9828 | 1 |
| contacts_age_grp2_infected | numeric | Contacts: Number of infected contacts who were within age group 2 | 9665 | 2 |
| contacts_age_grp3 | character | Contacts: Definition of age group 3, e.g, 25–64 yrs | 9799 | 1 |
| contacts_age_grp3_total | numeric | Contacts: Number of contacts who were within age group 3 | 9871 | 0 |
| contacts_age_grp3_infected | numeric | Contacts: Number of infected contacts who were within age group 3 | 9809 | 1 |
| contacts_age_grp4 | character | Contacts: Definition of age group 4, e.g, >= 65 years old. | 9816 | 1 |
| contacts_age_grp4_total | numeric | Contacts: Number of contacts who were within age group 4 | 9877 | 0 |
| contacts_age_grp4_infected | numeric | Contacts: Number of infected contacts who were within age group 4 | 9824 | 1 |
| contacts_female | numeric | Contacts: Number of female contacts. | 9831 | 1 |
| contacts_female_infected | numeric | Contacts: Number of infected female contacts. | 9294 | 6 |
| contacts_male | numeric | Contacts: Number of male contacts. | 9832 | 1 |
| contacts_male_infected | numeric | Contacts: Number of infected male contacts. | 9299 | 6 |
| contacts_num_vaccinated | numeric | Contacts: number of fully vaccinated contacts. | 9875 | 0 |
| contacts_num_vaccinated_infected | numeric | Contacts: number of infected fully vaccinated contacts. | 9874 | 0 |
| contacts_context | character | Contacts: context. Describe the setting of each contact. | 4886 | 51 |
| contacts_infected_context | character | Contacts: context. Describe the setting of each infected contact. | 8034 | 19 |
| variant | character | Variant name | 0 | 100 |
| prevention_context | character | Prevention context | 7413 | 25 |

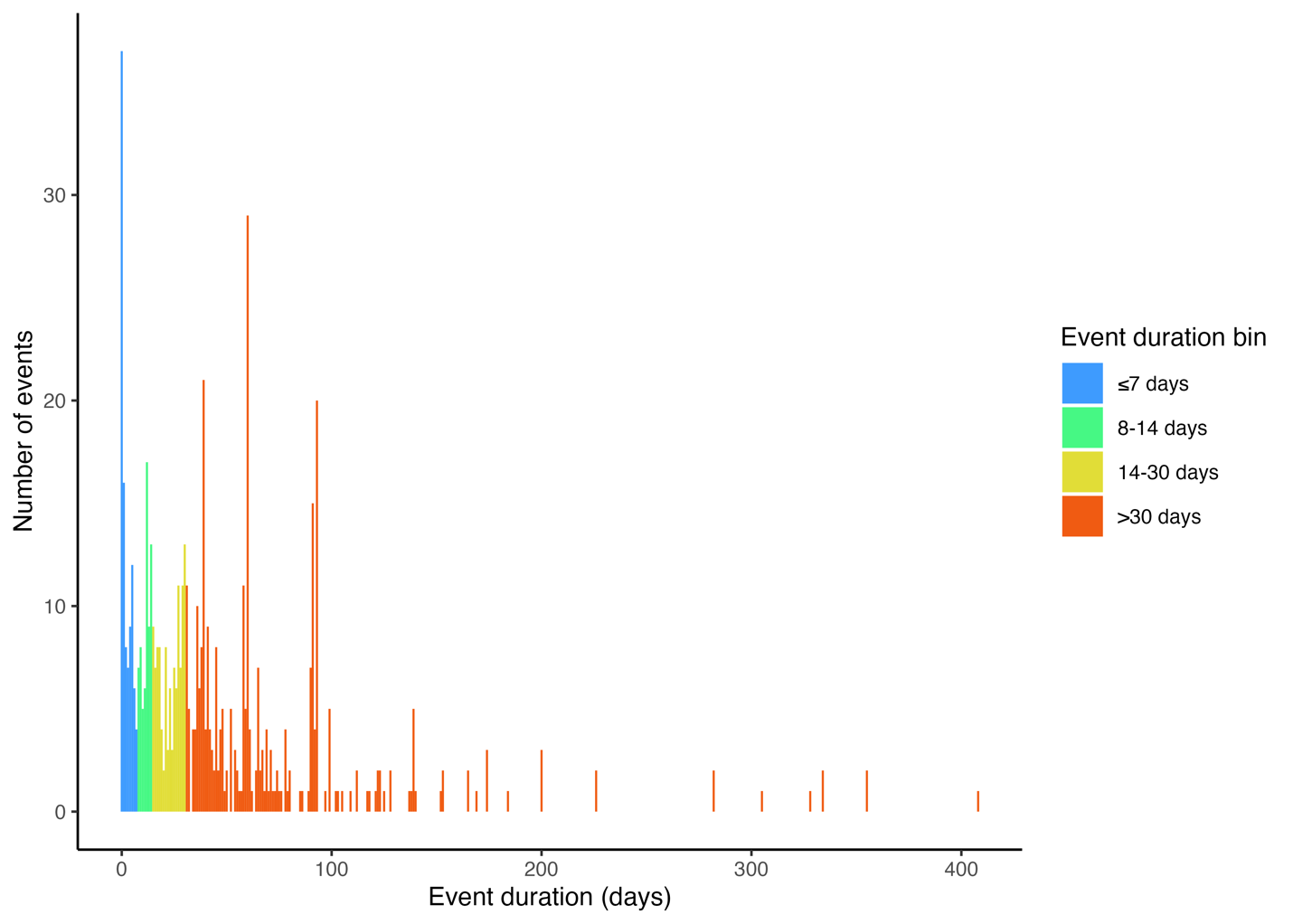

**Supplementary Figure S3.** Reported duration of transmission events reported in papers reviewed from December 2019 to August 2021. Event durations were calculated as the difference between the reported start and end of the exposure window for the transmission event.

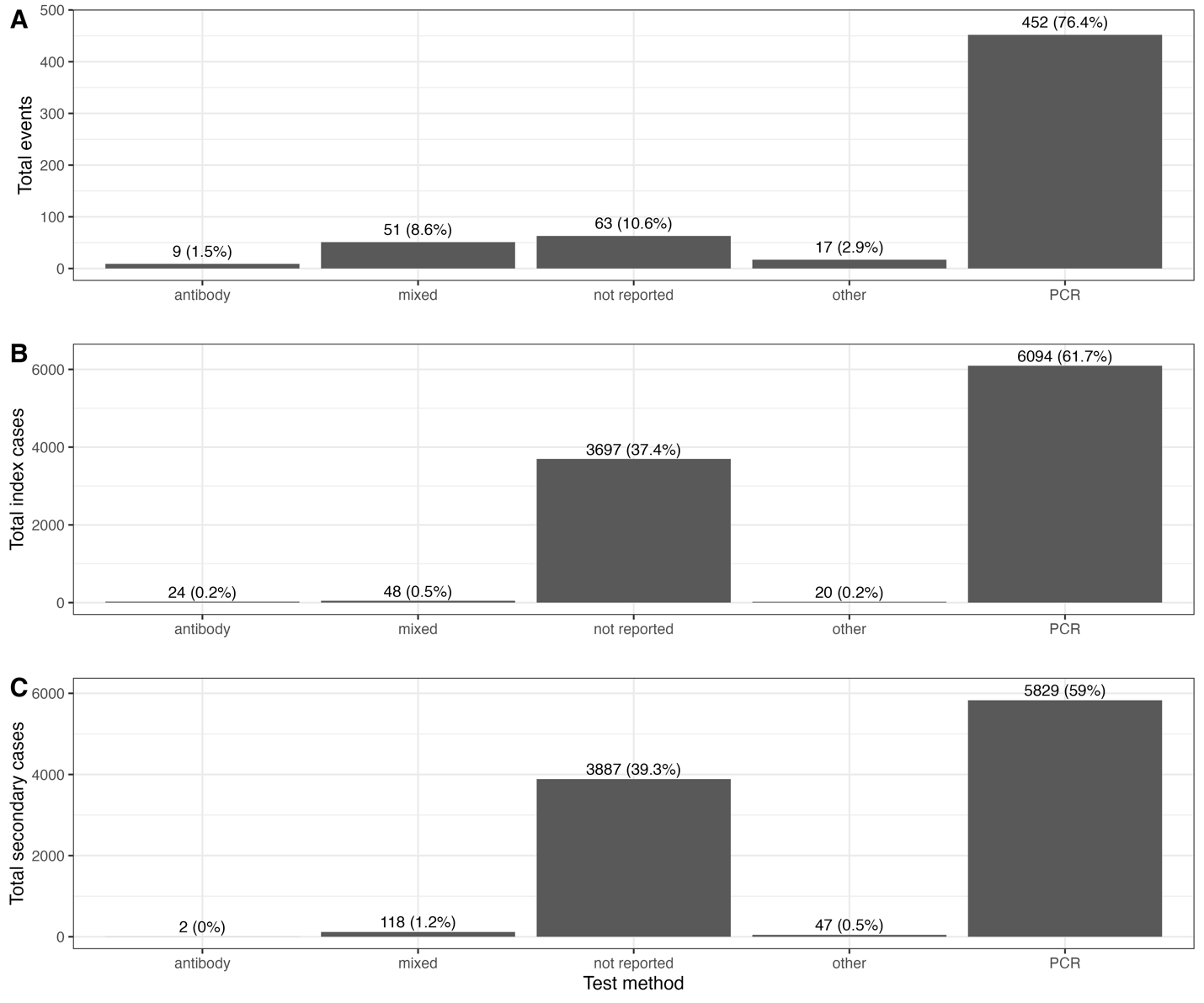

**Supplementary Figure S4.** Diagnostic methods for identification of SARS-CoV-2 cases across papers reviewed from December 2019 to August 2021 for transmission events (A), individual index cases (B), and secondary cases associated with an individual index case (C). “Other” methods included diagnosis based on key COVID-19 symptoms, abnormal chest tomography scans, or antigen tests. “Mixed” included any combination of PCR, antibodies, or other methods.

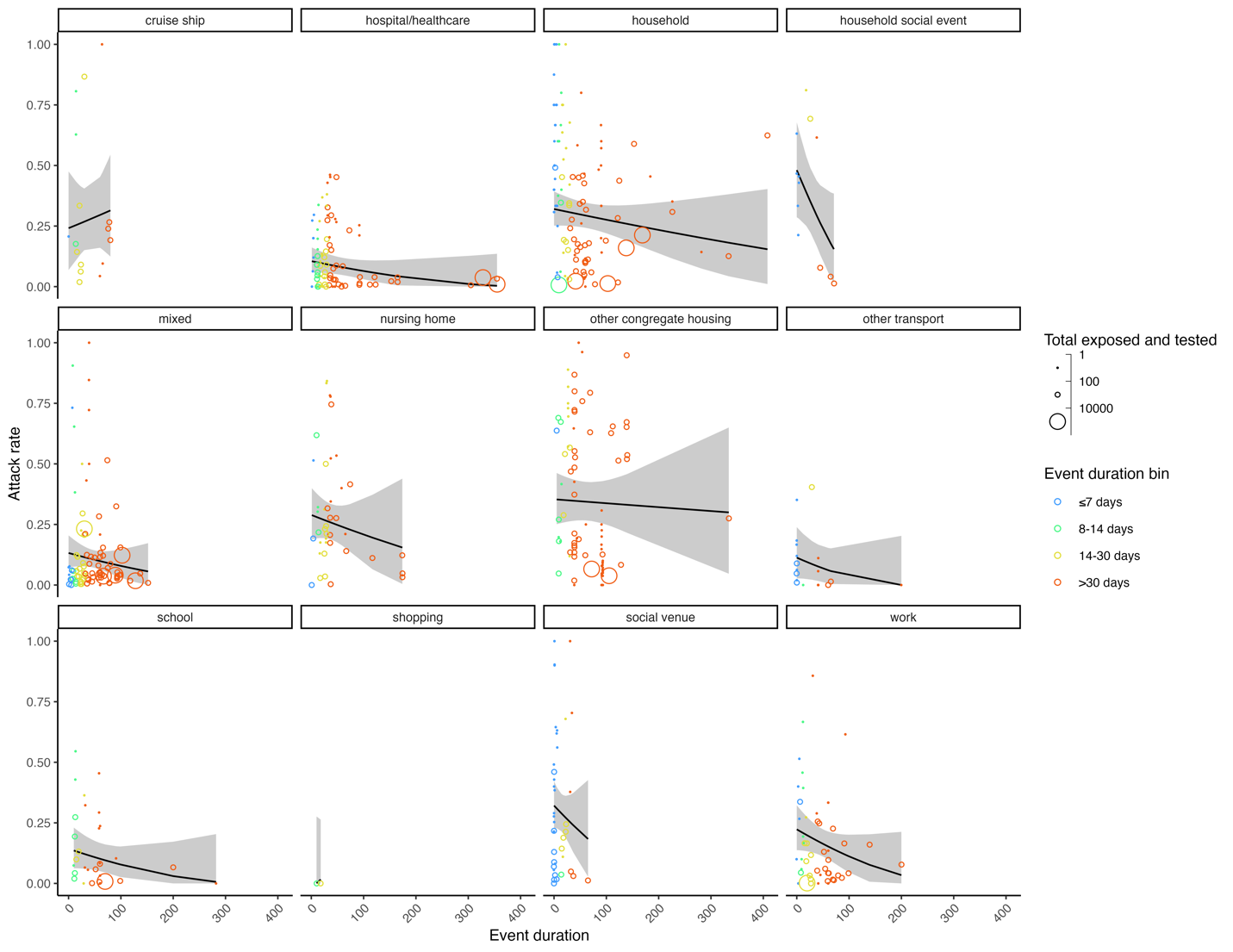

**Supplementary Figure S5.** The relationship between SARS-CoV-2 secondary attack rates and continuous event duration across 12 event types. Data included events occurring between December 2019 and August 2021 reported in the literature across 592 events from 296 studies. Individual event secondary attack rates are shown as bubbles colored by binned event duration, varying in size according to the total number of individuals exposed and tested from the event. Meta-analysis estimated relationships between secondary attack rates and event duration for each event type are shown as black lines along with the estimated 95% confidence intervals.

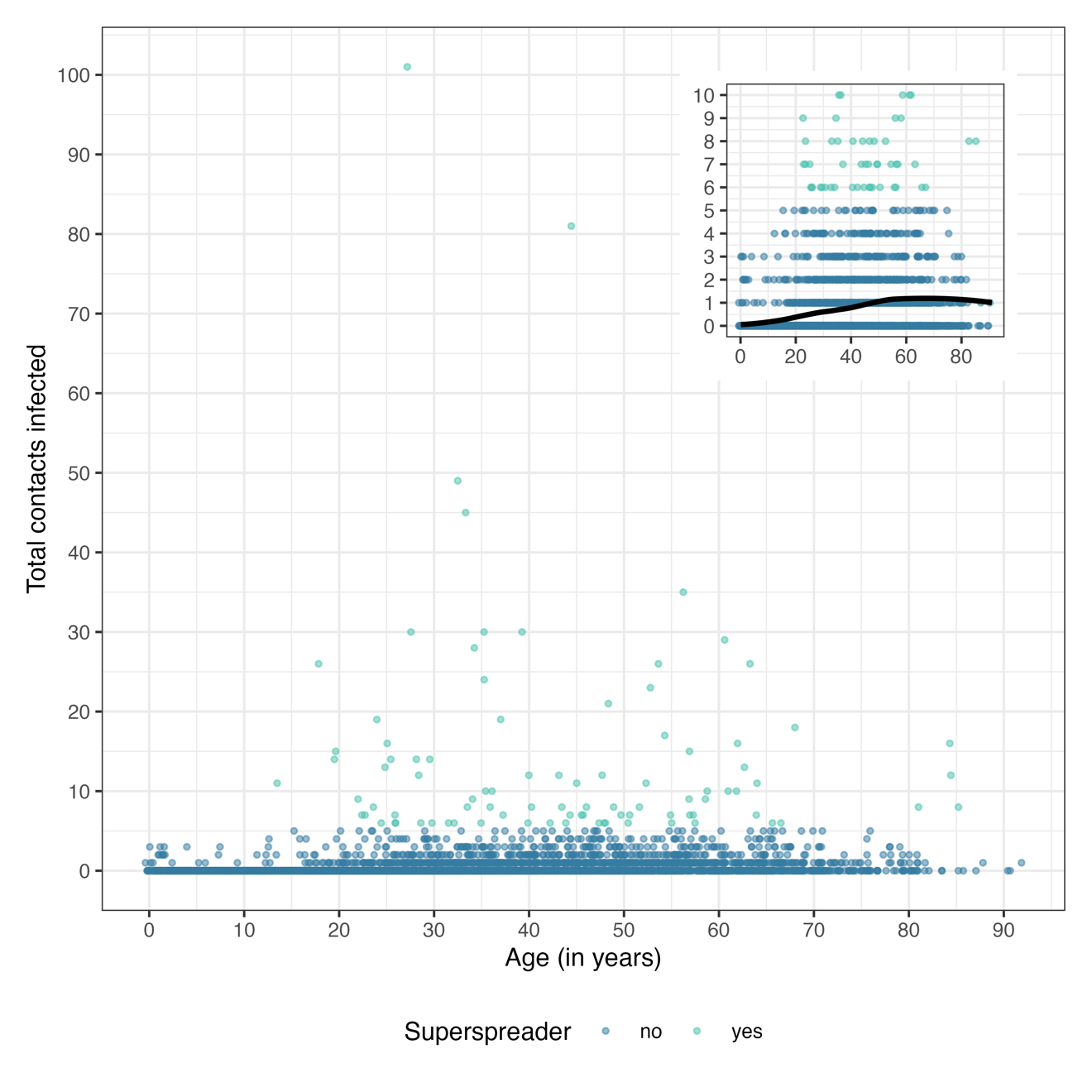

**Supplementary Figure S6.** Relationship between the number of secondary contacts infected by individual index cases and index case age (N = 9,591) for SARS-CoV-2 cases occurring between December 2019 and July 2021 reported in 259 studies. The inset shows a portion of the same data to highlight the distribution of superspreaders (index cases with >5 secondary cases). The black line in the inset displays the locally estimated scatterplot smoothing function for the points.

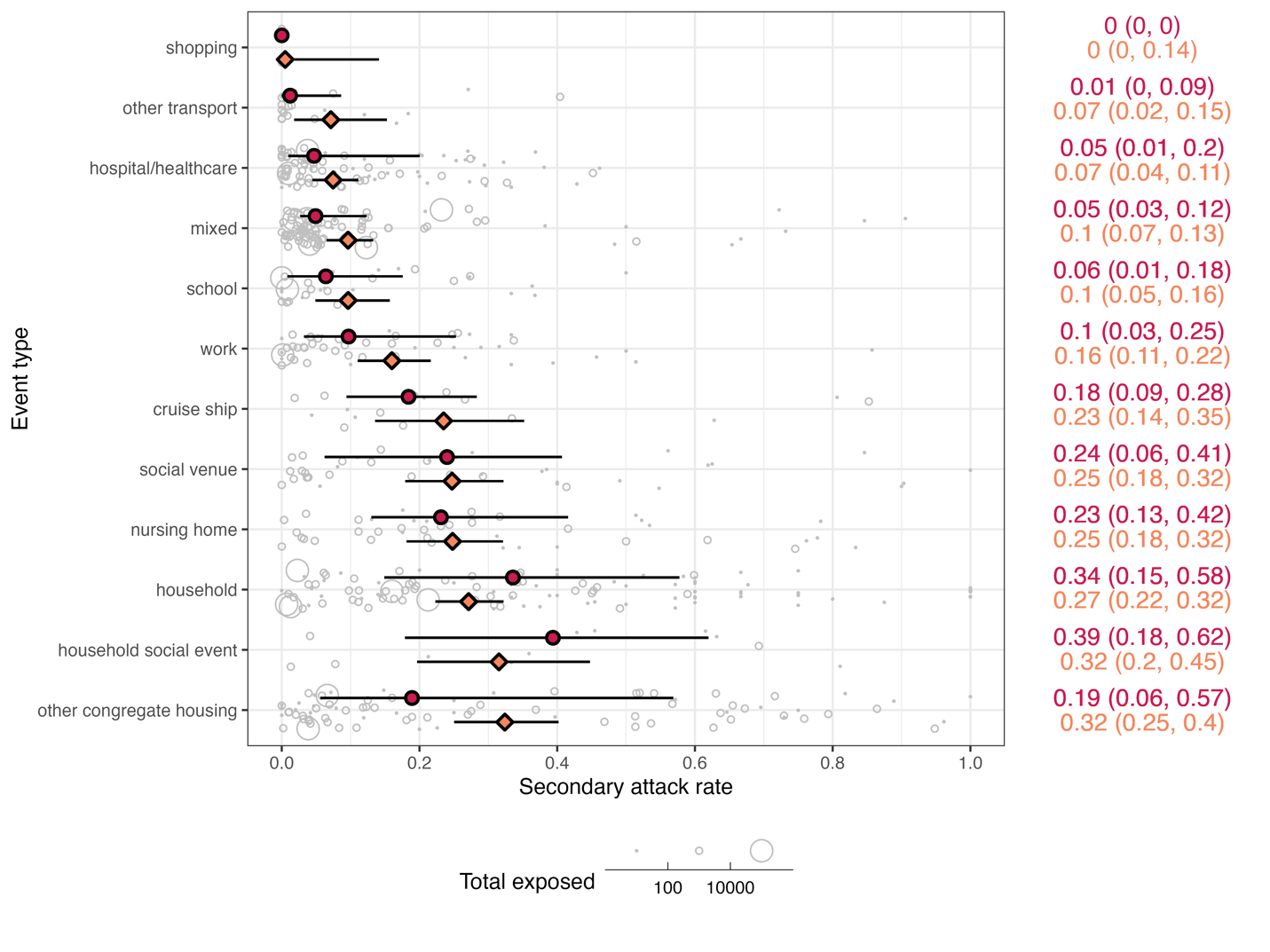

**Supplementary Figure S7.** SARS-CoV-2 secondary attack rates across 12 event types occurring between December 2019 and August 2021 reported in the literature across 592 events from 296 studies (complement to Figure 2). Individual event data secondary attack rates are shown as grey bubbles, varying in size according to the total number of individuals exposed from the event. Median secondary attack rate for each event type is shown as red circle with a line representing the interquartile range; values are in red on the right side of the figure. Meta-analysis estimated secondary attack rate for each event type is shown as an orange diamond with a line representing the estimated 95% confidence interval; values are in orange on the right side of the figure. Event types were ranked by increasing estimated mean SAR along the left axis. *I^2^*_total_ = 99%, *I^2^*_study_ = 58%, *I^2^*_event_ = 41%; Cochran’s *Q_E_*_,590_ = 153525, *P* < 0.0001; Cochran’s *Q_M_*_,11_ = 121, *P* < 0.0001.

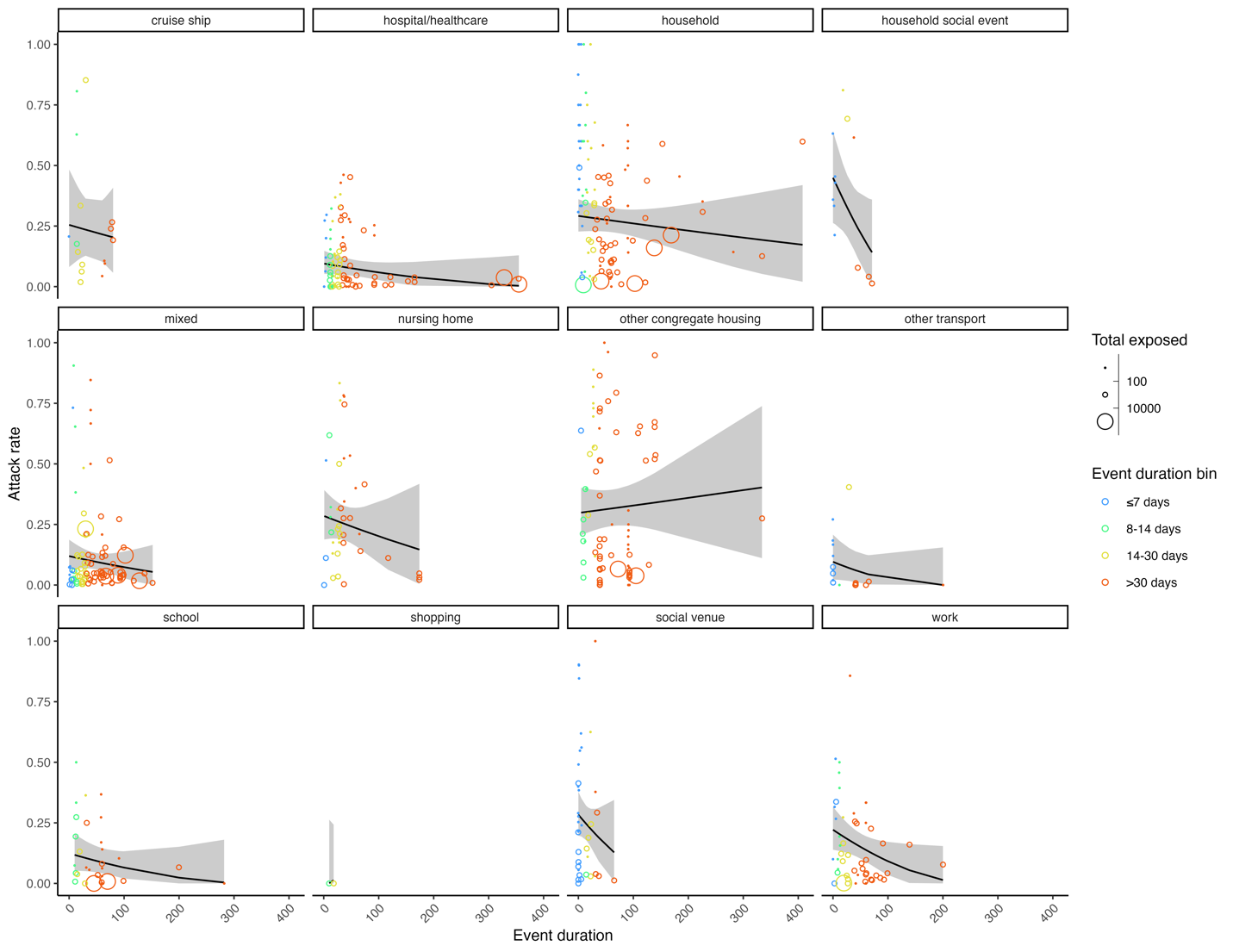

**Supplementary Figure S8.** The relationship between SARS-CoV-2 secondary attack rates and continuous event duration across 12 event types (complement to Supplementary Figure S5). Data included events occurring between December 2019 and August 2021 reported in the literature across 592 events from 296 studies. Individual event secondary attack rates are shown as bubbles colored by binned event duration, varying in size according to the total number of individuals exposed from the event. Meta-analysis estimated relationships between secondary attack rates and event duration for each event type are shown as black lines along with the estimated 95% confidence intervals. Cochran’s *Q_E_*_,567_ = 85563, *P* < 0.0001; Cochran’s *Q_M_*_,23_ = 136, *P* < 0.0001.

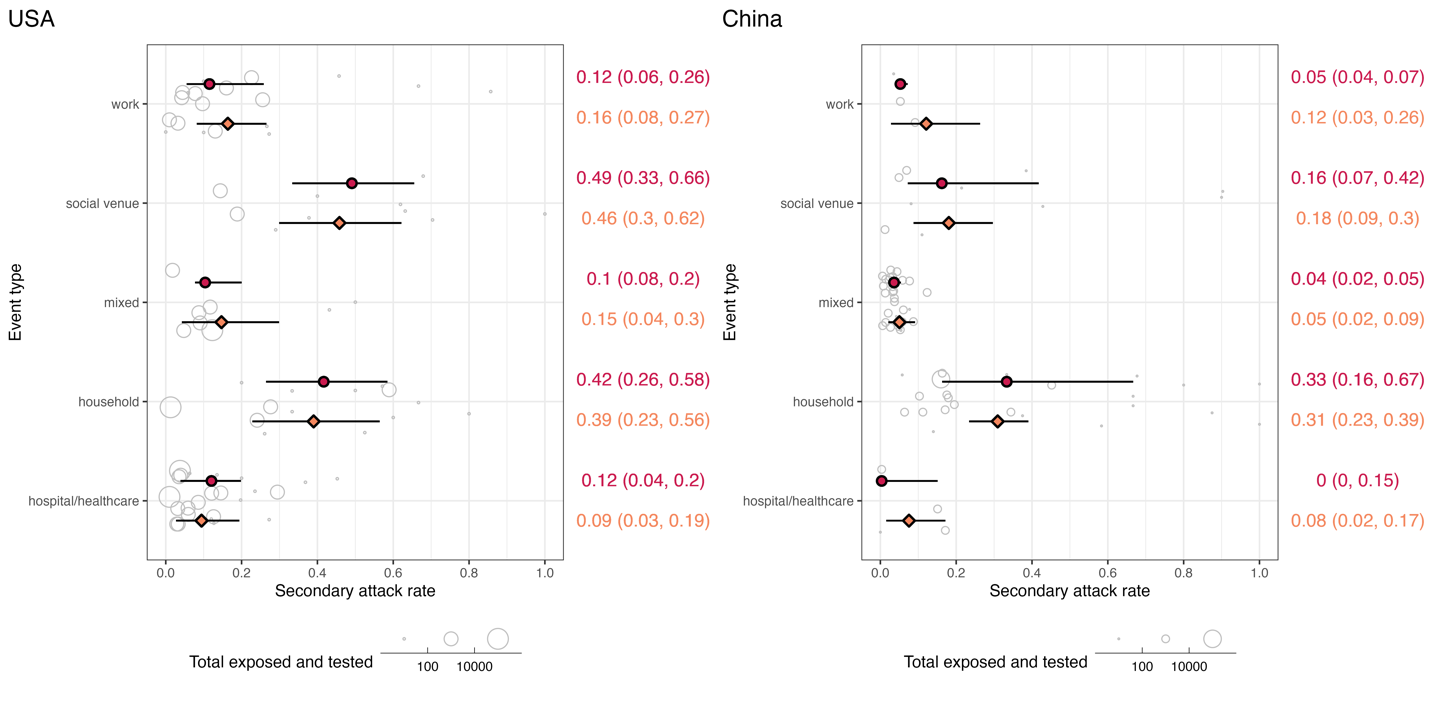

**Supplementary Figure S9.** SARS-CoV-2 secondary attack rates across 12 event types occurring between December 2019 and August 2021 reported in the literature from the USA and China. Comparison by event type across the two countries was only possible for a subset of five event types with sufficient data (i.e., more than one event for an event type in each country). Individual event data secondary attack rates are shown as grey bubbles, varying in size according to the total number of individuals exposed and tested from the event. Median secondary attack rate for each event type is shown as red circle with a line representing the interquartile range; values are in red on the right side of the figure. Meta-analysis estimated secondary attack rate for each event type is shown as an orange diamond with a line representing the estimated 95% confidence interval; values are in orange on the right side of the figure.

**Supplementary Table S5.** Results of sensitivity analysis “loops” among SARS-CoV-2 index cases. Total rows of data included in each comparison are shown for each strategy.

| **Statistical test** | **Original strategy: drop loops only (N = 9,591)** | **Alternative strategy 1: keep loops (N = 9,883)** | **Alternative strategy 2: remove all index cases in studies with loops (N = 8,149)** | **Alternative strategy 3: keep loops but randomly draw values from a binomial distribution (N = 9,883)** |
| --- | --- | --- | --- | --- |
| Negative binomial parameters | mean: 0.88 (CI 0.84–0.92) *k*: 0.27 (CI 0.25–0.28) | mean: 0.94 (CI: 0.9–0.98)  *k*: 0.29 (CI 0.28–0.31) | mean: 0.87 (CI: 0.82–0.91)  *k*: 0.22 (CI 0.21–0.23) | average mean over 100 draws: 0.92  average *k* over 100 draws: 0.28 |
| Chi-square test of proportion women among superspreaders vs. non-superspreaders | χ^2^_1_ = 0.09, *P* = 0.76 | χ^2^_1_ = 0.008, *P* = 0.93 | χ^2^_1_ = 0.25, *P* = 0.62 | all 100 *P* ≥ 0.05 |
| Chi-square test of proportion symptomatic among superspreaders vs. non-superspreaders | χ^2^_1_ = 5.4, *P* = 0.02 | χ^2^_1_ = 6.5, *P* = 0.01 | χ^2^_1_ = 6.6, *P* = 0.01 | all 100 *P* < 0.05 |
| Chi-square test of proportion of age (7 age bins) among superspreaders vs. non-superspreaders | χ^2^_6_ = 21.7, *P* = 0.001 | χ^2^_6_ = 22, *P* = 0.001 | χ^2^_6_ = 20.9, *P* = 0.002 | all 100 *P* < 0.005 |
| Chi-square test of proportion of age (≥18 years) among superspreaders vs. non-superspreaders | χ^2^_1_ = 14.1, *P* < 0.0001 | χ^2^_1_ = 13.5, *P* = 0.0002 | χ^2^_1_ = 12.5, *P* = 0.0003 | all 100 *P* < 0.0005 |
| t-test of age (in years) among superspreaders vs. non-superspreaders | t_94.4_ = 5.2, *P* < 0.0001 | t_105_ = 5.4, *P* < 0.0001 | t_84.8_ = 4.7, *P* < 0.0001 | all 100 *P* < 0.0001 |
| t-test of Ct value among superspreaders vs. non-superspreaders | t_2.69_ = -0.5, *P* = 0.62 | t_2.55_ = -0.45, *P* = 0.69 | t_5.09_ = -0.1, *P* = 0.92 | all 100 *P* ≥ 0.05 |
| Kruskal-Wallis test of total contacts among superspreaders vs. non-superspreaders | χ^2^_1_ = 56.6, *P* < 0.0001 | χ^2^_1_ = 58.4, *P* < 0.0001 | χ^2^_1_ = 53.2, *P* < 0.0001 | all 100 *P* < 0.0001 |

**Supplementary Table S6.** Statistical comparisons of asymptomatic versus symptomatic SARS-CoV-2 cases based on features reported in the literature in 259 studies for cases occurring between December 2019 and July 2021.

| **Feature of comparison** | **Percentage or estimated mean for asymptomatic index cases (total observations)** | **Percentage or estimated mean for symptomatic index cases (total observations)** | **Statistical test results** |
| --- | --- | --- | --- |
| Female | 50% (N = 66) | 46% (N = 439) | χ^2^_1_ = 0.26, *P* = 0.61 |
| Age (in bins)  ≤4 years  5–12 years  13–18 years  19–24 years  25–48 years  49–64 years  ≥65 years | (N = 62)  2%  5%  2%  6%  56%  21%  8% | (N = 400)  2%  2%  2%  6%  45%  30%  13% | χ^2^_6_ = 5.6, *P* = 0.47 |
| Age (≥18 years) | 92% (N = 62) | 93% (N = 400) | χ^2^_1_ = 0.03, *P* = 0.85 |
| Age (in years) | 38.8 (N = 66) | 44.4 (N = 448) | t_82.1_ = 2.3, *P* = 0.02 |
| Ct value | 30.2 (N = 12) | 28.4 (N = 58) | t_17.1_ = 0.97, *P* = 0.35 |
| Total contacts | 58 (N = 28) | 56 (N = 171) | χ^2^_1_ = 0.38, *P* = 0.54 |
